# Proteomic studies of human placentas reveal partnerships associated with preeclampsia, diabetes, gravidity, and labor

**DOI:** 10.1101/2023.07.23.23292673

**Authors:** Shannon J. Ho, Dale Chaput, Rachel G. Sinkey, Amanda H. Garces, Erika P. New, Maja Okuka, Peng Sang, Sefa Arlier, Nihan Semerci, Thora S. Steffensen, Thomas J. Rutherford, Angel E. Alsina, Jianfeng Cai, Matthew L. Anderson, Ronald R. Magness, Vladimir N. Uversky, Derek A. T. Cummings, John C. M. Tsibris

**Affiliations:** Department of Obstetrics and Gynecology, University of South Florida, Tampa, Florida, USA; Department of Cell Biology, Microbiology and Molecular Biology, University of South Florida, Tampa, Florida, USA; Lisa Muma Weitz Microscopy Laboratory, University of South Florida, USA; Department of Chemistry, University of South Florida, Tampa, Florida, USA; Department of Pathology, Tampa General Hospital, Tampa, Florida, USA; Cancer Center, Tampa General Hospital, Tampa, Florida, USA; Transplant Surgery Center, Tampa General Hospital, Tampa, Florida, USA; Department of Molecular Medicine, University of South Florida, Tampa, Florida, USA; Department of Biology and Emerging Pathogens Institute, University of Florida, Gainesville, FL, USA

## Abstract

VEGFR2 is a central regulator of placental angiogenesis. The study of the VEGFR2 proteome of chorionic villi at term revealed its partners MDMX and PICALM. The oxytocin receptor (OT-R) and vasopressin V1aR receptor were detected in MDMX and PICALM immunoprecipitations. Immunogold electron microscopy showed VEGFR2 on endothelial cell (EC) nuclei, mitochondria, and Hofbauer cells (HC), the tissue-resident macrophages. MDMX, PICALM, and V1aR were on EC plasma membranes, nuclei, and HC nuclei. Unexpectedly, PICALM and OT-R were detected on EC projections into the fetal lumen and OT-R on 20-150 nm clusters therein, prompting the hypothesis that placental exosomes transport OT-R to the fetus and across the blood-brain barrier. Insights on gestational complications were gained by univariable and multivariable regression analyses associating preeclampsia with lower MDMX protein levels in membrane extracts of chorionic villi, and lower MDMX, PICALM, OT-R, and V1aR with spontaneous vaginal deliveries compared to cesarean deliveries before labor. We found select associations between higher MDMX, PICALM, OT-R and either gravidity, diabetes, BMI, maternal age, or neonatal weight, and correlations between PICALM-OT-R (p<2.7x10^-8^), PICALM-V1aR (p<0.006), and OT-R-V1aR (p<0.001). These results offer for exploration new partnerships in metabolic networks, tissue-resident immunity, and labor, notably for HC that predominantly express MDMX.

---

The placenta is a transient organ that performs the functions of major organs of the fetus, such as lungs, liver, and kidney^1^, supplies the fetus and in particular the fetal brain with oxygen and nutrients, and facilitates waste disposal and provides immune protection^2, 3^. The genomes of placenta and fetus are identical except in cases of confined placental mosaicism. The vascular endothelial growth factor A (VEGF-A) is a key regulator of vasculogenesis, angiogenesis and placental growth that acts mainly through VEGFR1 and VEGFR2, two tyrosine kinase single-pass transmembrane receptors^4–6^.

Preeclampsia is a serious complication of human pregnancy occurring in 5-7% of all gestations with newly-onset hypertension and proteinuria as its primary clinical characteristics^2, 7^. Preeclampsia is a multisystemic syndrome of different subtypes associated with serious health problems to mother and child even after pregnancy^8^. Lipid bilayer-enclosed extracellular vehicles (EV) transport extracellular nucleic acids, proteins, lipids, and metabolites^9–12^. Cancer cells deploy EV to activate VEGF signaling in endothelial cells^12^. Exosomes are EV measuring 20–150 nm in diameter. Placental-derived exosomes released in the maternal circulation are associated with pregnancy disorders and parturition^11^.

Tissue-based maps of the human proteome of many organs, including placentas, have been published,^13–16^ but great challenges remain to discover which among the hundreds of detected proteins regulate key metabolic networks, specifically, during normal and complicated pregnancies and labor. To obtain such information, we chose to immunoprecipitate VEGFR2, an ideal target as an extensively documented regulator of placental angiogenesis. To deploy a wide net for the membrane partners of VEGFR2, we analyzed the pellets obtained after high-speed centrifugation of chorionic villi homogenates. Uterine blood in the maternal intervillous space exchanges substances with fetal blood at the villous tree. Although the villi are physically separated from uterine blood^1, 3^, the pellets contain cells and EV from it. To preserve protein complexes, we extracted the pellets with ASB-14 (amidosulfobetaine-14), an efficient non-denaturing detergent. The extracts were immunoprecipitated with the bait antibody (Ab) charged on magnetic beads knowing that extraction of membrane proteins from their native environment could alter their structure and protein links. Mass spectrometry identified proteins immunoprecipitated with VEGFR2, especially its newly discovered placental partners, the multifunctional MDMX (Double minute 4 protein) and PICALM (Phosphatidylinositol-binding clathrin assembly protein).

MDMX, also known as MDM4 and HDMX, is a zinc-binding protein and a p53 inhibitor acting in coordination with MDM2, a zinc-dependent E3 ubiquitin ligase^17^. MDMX and MDM2 have numerous p53-independent activities. In preeclampsia, p53 is upregulated in villous trophoblasts^18^. PICALM is a nuclear and plasma membrane protein that interacts with phosphatidylinositol to recruit clathrin and adaptor protein-2, initiates endocytosis of clathrin-coated vesicles, internalizes ligand-receptor complexes, and participates in iron and cholesterol homeostasis^19, 20^. PICALM is a genetic risk factor for late-onset Alzheimer’s disease that participates in amyloid-β transcytosis and processing of amyloid precursor protein (APP). Human placentas express APP and APP-processing enzymes which are increased in preeclampsia^21^. Oxytocin is a hydrophilic neuropeptide and Pitocin, a synthetic oxytocin, is prescribed in the USA to induce labor and decrease postpartum hemorrhage. The oxytocin receptor (OT-R, *OXTR*) is a magnesium-dependent G protein-coupled receptor^22^ that activates a phosphatidylinositol-calcium second messenger system^23^. OT-R participates in numerous activities ranging from parturition to lactation and mother-child bonding^22, 23^. OT-R functions as a homodimer and in heterocomplexes with vasopressin receptors V1aR (*AVPR1A)*^24^, V2R, and other receptors. V1aR is the most abundant among vasoactive receptors in human arteries^25^.

Here, among the newly detected partners of VEGFR2 we focused on MDMX and PICLAM based on their documented biochemical functions, and their partners OT-R and V1aR. Finding OT-R in exosome-size clusters in the fetal lumen led to the hypothesis that placental OT-R is carried to the fetus in exosomes, potential transporters of future therapeutic agents. MDMX, PICALM, OT-R, and V1aR protein levels, estimated by western blots and relative to an internal control sample, were associated with clinical characteristics of the 44 patients we studied. Potential insights on the molecular mechanisms of placental MDMX and PICALM were gained, respectively, from the cancer and Alzheimer’s disease literature.

## Results

### Immunoprecipitations (IP)

Key prerequisite for this study was that VEGFR2 protein complexes withstood tissue freezing and extraction by ASB-14. Since Blue-Native electrophoresis (BN) resolves protein mixtures under non-denaturing conditions, we fractionated placental extracts by BN followed by VEGFR2 immunostaining which revealed streaks extending up to 800 kDa. VEGFR2 monomers appear at 220 kDa in western blots. Apparently, ASB-14 stabilized the extracted protein complexes of VEGFR2. Several proteins described in this study were efficiently extracted by ASB-14 (see Methods). New VEGFR2 partners, MDMX and PICALM, were also selected for IP.

Fig. 1 shows western blots of the fractions eluted from the magnetic beads and used to select the fraction for mass spectrometric analysis. Representative fractions are identified by the patient’s assigned letter-number code shown on Table 1. Magnetic beads retained all of the applied VEGFR2 (**A**) as none was detected in the flow-through fraction, but the MDMX and PICALM antibodies charged on the beads retained only a fraction of their target protein as shown, respectively, in **E** and **F**. All target proteins were greatly enriched in the eluted fraction-E relative to the applied placental extract, fraction-X. OT-R co-immunoprecipitated with MDMX (**G**, right side) and with PICALM (**G**, left side), but not with VEGFR2 (**D**). V1aR co-immunoprecipitated with MDMX (**H**, left side) and with PICALM (**H**, right side), but not with VEGFR2 (**C**). Eluted MDMX appeared smaller in the VEGFR2 IP (**B**) and MDMX IP (**E**), whereas PICALM (**F**), OT-R (**G**) and V1aR (**H**) appeared larger. The size of eluted VEGFR2 was the same as in the extracts (**A**).

**Fig. 1.**
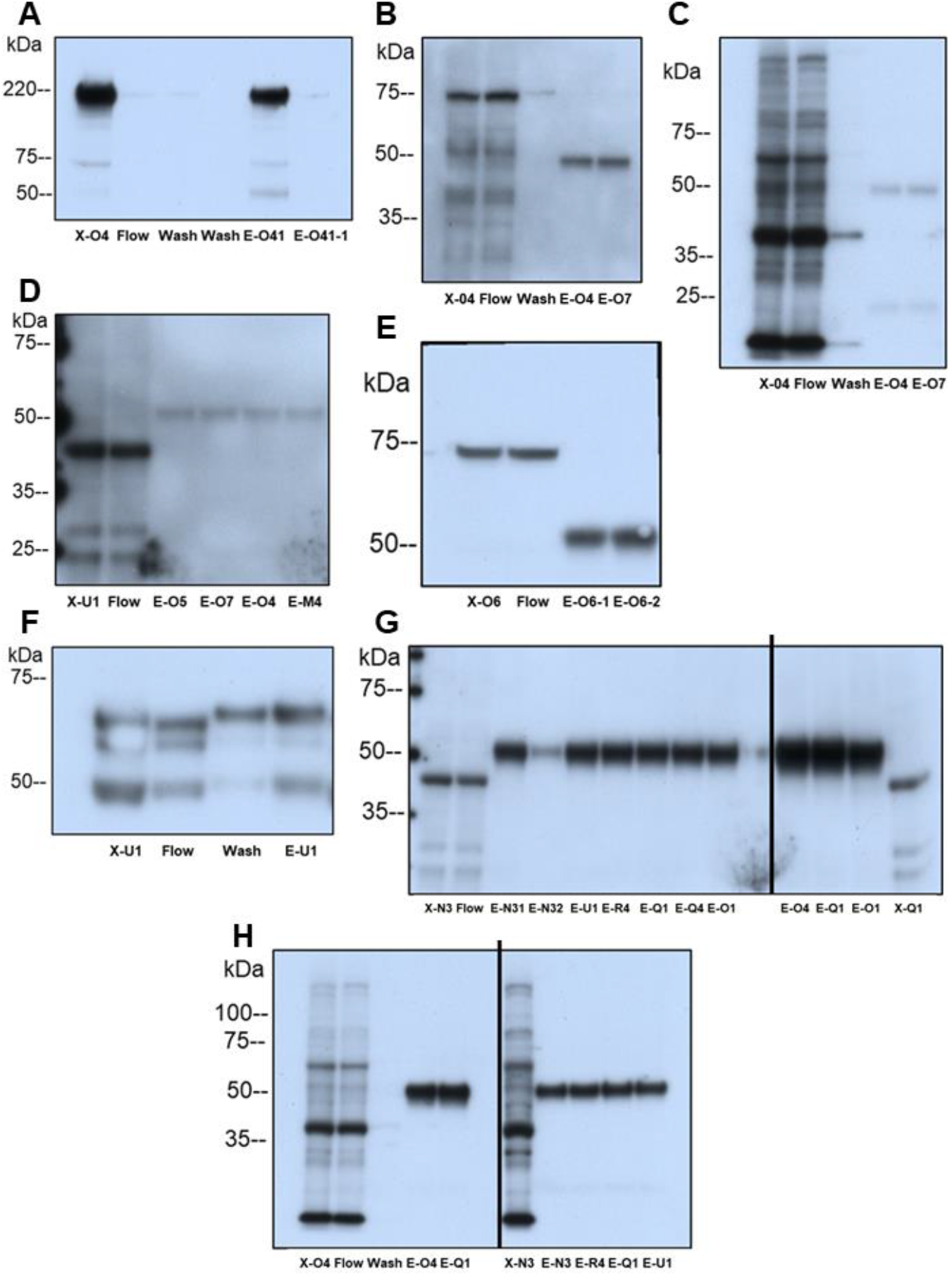
**Protein elution profiles in VEGFR2, MDMX and PICALM IP**. Detergent-extracted villous sample X was incubated with magnetic bead suspension charged with the target Ab (see Methods). E denotes eluted protein fractions. Western blots detected the target proteins eluted in the uniformly performed IP experiments. **A.** VEGFR2 in VEGFR2-IP. **B.** MDMX in VEGFR2-IP. **C.** V1aR in VEGFR2-IP. **D.** OT-R in VEGFR2-IP. **E.** MDMX in MDMX-IP. **F.** PICALM in PICALM-IP. **G.** Left-side, OT-R in samples from PICALM-IP, and right-side OT-R in samples from MDMX-IP. **H.** Left-side, V1aR in samples from MDMX-IP, and right-side samples from PICALM-IP. Panel **C** shows the membrane in **B** reprobed for V1aR without stripping.

**Table 1.** Clinical characteristics of 47 patients in the study group. A. Patient samples were labelled with a code consisting of a letter and 1, 2, 5, 6, 7 for non-preeclamptic, 3, 4 for preeclamptic patients or a Ho-number**. B**. Patients Ho-71, Ho- 72 and Ho-73 provided placental samples for immunogold electron microscopy.

| A |  |  |  |  |  |  |  |  |  |  |  |  |
| --- | --- | --- | --- | --- | --- | --- | --- | --- | --- | --- | --- | --- |
| Patient | Age Range | Gravida | Gestational Age (weeks) | BMI in L&D | Race | cHTN | Preeclampsia | DM | Delivery Mode | Pitocin pre-delivery | Sex F=1 | Neonatal weight (g) |
| H1 | 21-25 | 1 | 38.0 | 30.0 | 6 | 0 | 0 | 0 | 1 | 1 | 1 | 2,940 |
| H2 | 21-25 | 2 | 37.0 | 24.0 | 8 | 0 | 0 | 0 | 1 | NA | 2 | 2,590 |
| H3 | 31-35 | 2 | 37.0 | 32.0 | 8 | 0 | 1 | 0 | 1 | NA | 1 | 3,040 |
| I-1 | 21-25 | 3 | 38.0 | 34.0 | 2 | 0 | 0 | 0 | 3 | 1 | 2 | 2,631 |
| I-2 | 21-25 | 1 | 42.0 | 30.0 | 1 | 0 | 0 | 0 | 1 | 1 | 2 | 3,759 |
| I-3 | 26-30 | 8 | 35.0 | 34.0 | 2 | 0 | 1 | 2 | 2 | 0 | 2 | 3,195 |
| I-4 | 18-20 | 1 | 36.6 | 41.0 | 8 | 0 | 1 | 0 | 3 | 0 | 1 | 3,335 |
| J-1 | 26-30 | 2 | 39.0 | 25.0 | 8 | 0 | 0 | 0 | 3 | 0 | 2 | 3,300 |
| J-2 | 36-40 | 2 | 37.4 | 22.0 | 1 | 0 | 0 | 0 | 1 | 1 | 1 | 3,460 |
| J-3 | 26-30 | 3 | 38.5 | 28.0 | 1 | 0 | 1 | 3 | 3 | 1 | 2 | 3,630 |
| L1 | 21-25 | 1 | 39.0 | 24.0 | 3 | 0 | 0 | 0 | 3 | 0 | 1 | 3,147 |
| L2 | 18-20 | 1 | 39.0 | 29.0 | 6 | 0 | 0 | 0 | 1 | 1 | 1 | 2,920 |
| L3 | 21-25 | 3 | 35.0 | 30.0 | 2 | 0 | 1 | 0 | 2 | 0 | 2 | 2,235 |
| L4 | 26-30 | 3 | 40.0 | 29.0 | 8 | 0 | 1 | 0 | 1 | NA | 1 | 3,010 |
| M1 | 21-25 | 3 | 40.1 | 55.0 | 2 | 0 | 0 | 0 | 2 | 0 | 1 | 3,795 |
| M3 | 18-20 | 2 | 37.0 | 31.3 | 8 | 0 | 1 | 0 | 2 | 0 | 1 | 3,490 |
| M4 | 26-30 | 1 | 40.1 | 39.8 | 8 | 0 | 1 | 0 | 1 | 1 | 2 | 3,380 |
| N1 | 31-35 | 4 | 39.1 | 30.0 | 8 | 0 | 0 | 2 | 2 | 0 | 2 | 3,800 |
| N2 | 31-35 | 2 | 39.3 | 32.8 | 8 | 0 | 0 | 0 | 2 | 0 | 1 | 3,180 |
| N3 | 31-35 | 2 | 39.0 | 35.5 | 8 | 0 | 1 | 0 | 2 | 0 | 2 | 3,390 |
| N4 | 18-20 | 2 | 36.4 | 29.5 | 1 | 0 | 1 | 0 | 3 | 1 | 2 | 3,510 |
| O1 | 21-25 | 2 | 39.3 | 26.2 | 1 | 0 | 0 | 0 | 4 | 1 | 1 | 2,960 |
| O2 | 21-25 | 1 | 38.4 | 34.8 | 1 | 0 | 0 | 0 | 1 | 1 | 1 | 2,940 |
| O3 | 26-30 | 1 | 37.1 | 29.5 | 1 | 0 | 1 | 0 | 1 | 1 | 1 | 2,800 |
| O4 | 21-25 | 3 | 40.3 | 52.5 | 1 | 0 | 1 | 0 | 3 | 1 | 2 | 3,575 |
| O5 | 36-40 | 4 | 39.1 | 50.4 | 8 | 0 | 0 | 3 | 2 | 0 | 2 | 4,295 |
| O7 | 32-35 | 12 | 39.3 | 35.2 | 2 | 0 | 0 | 0 | 2 | 0 | 1 | 3,780 |
| Q1 | 21-25 | 2 | 37.0 | 47.8 | 1 | 0 | 0 | 3 | 2 | 0 | 1 | 3,885 |
| Q2 | 31-35 | 4 | 39.1 | 35.8 | 2 | 0 | 0 | 3 | 2 | 0 | 1 | 3,390 |
| Q3 | 26-30 | 2 | 37.2 | 66.7 | 1 | 1 | 1 | 0 | 3 | 1 | 2 | 3,375 |
| Q4 | 31-35 | 4 | 39.1 | 77.2 | 2 | 0 | 1 | 3 | 2 | 0 | 2 | 3,835 |
| R3 | 21-25 | 1 | 37.6 | 49.5 | 1 | 1 | 1 | 0 | 1 | 1 | 2 | 2,560 |
| R4 | 26-30 | 4 | 39.1 | 29.8 | 2 | 0 | 1 | 2 | 3 | 1 | 1 | 3,960 |
| R5 | 21-25 | 6 | 39.0 | 30.9 | 2 | 0 | 0 | 0 | 2 | 0 | 1 | 3,205 |
| S1 | 26-30 | 2 | 37.4 | 46.2 | 8 | 0 | 0 | 2 | 2 | 0 | 2 | 4,675 |
| S2 | 26-30 | 3 | 37.5 | 42.2 | 1 | 0 | 0 | 2 | 2 | 0 | 1 | 4,600 |
| T1 | 26-30 | 5 | 38.0 | 41.9 | 1 | 0 | 0 | 2 | 2 | 0 | 1 | 3,090 |
| T2 | 26-30 | 1 | 39.1 | 53.3 | 1 | 0 | 0 | 2 | 2 | 0 | 2 | 3,960 |
| U1 | 31-35 | 3 | 37.1 | 62.6 | 2 | 1 | 0 | 2 | 2 | 0 | 2 | 4,440 |
| U2 | 26-30 | 3 | 37.1 | 52.7 | 1 | 1 | 0 | 2 | 2 | 0 | 2 | 5,020 |
| V1 | 36-40 | 5 | 37.1 | 35.5 | 2 | 1 | 0 | 0 | 2 | 0 | 2 | 2,720 |
| V2 | 36-40 | 2 | 38.4 | 26.1 | 1 | 0 | 0 | 0 | 2 | 0 | 2 | 4,150 |
| W1 | 18-20 | 1 | 39.2 | 30.5 | 1 | 0 | 0 | 0 | 2 | 0 | 2 | 3,470 |
| W2 | 26-30 | 8 | 37.0 | 51.7 | 2 | 1 | 0 | 0 | 2 | 0 | 1 | 3,000 |
| B |  |  |  |  |  |  |  |  |  |  |  |  |
| Ho-71 | 31-35 | 6 | 37.1 | NA | 2 | 0 | 0 | 0 | 2 | 0 | 1 | 3,195 |
| Ho-72 | 26-30 | 1 | 39.3 | 29.4 | 2 |  | 0 | 3 | 2 | NA | 1 | 3,400 |
| Ho-73 | 26-30 | 1 | 37.0 | 29.1 | 8 | 0 | 1 | 0 | 1 | 1 | 2 | 2,780 |

Abbreviations: *Race*: White 1, Black 2, Asian 3, Hispanic 8, unknown 6. *Chronic hypertension*: cHTN 1. *Preeclampsia* 1. *Diabetes (DM)*: Type-1 1, Type-2 2, Gestational 3. *Delivery mode*: Spontaneous vaginal delivery (SVD) 1, Cesarean delivery (CD) prior to the onset of labor 2, CD after the onset of labor 3, Vaginal birth after CD (VBAC) 4. *Pitocin pre-delivery* 1. *Fetal sex*: Female 1, Male 2. *L&D*: Labor and delivery suite. *NA*, not available. The number of patients in each category were as follows: Diabetes: 30 no, Type-2 9, Gestational 5. Preeclampsia: 28 no, 16 yes. Delivery Mode: SVD 12; CD prior to the onset of labor 23; CD after the onset of labor 9; VBAC 1.

### Proteomic analysis

Results of the immunoprecipitations of VEGFR2 (samples O4, O5, O7, U1), MDMX (samples O1, O4, O6, Q1, Q4, R4), and PICALM (samples N3, O1, R4, Q1, Q4, U1) are listed in Tables 2-4. The peptide coverage was not consistent among the placental extracts in each immunoprecipitation group due to different clinical characteristics of the patients whose tissues we tested, and potential differences in post-translational modifications, such as acetylation, and ubiquitination. Soluble proteins in villous homogenates having an affinity for membrane proteins would be retained on the pellets obtained after centrifugation at 100,000*g* for 1h.

**Table 2.**
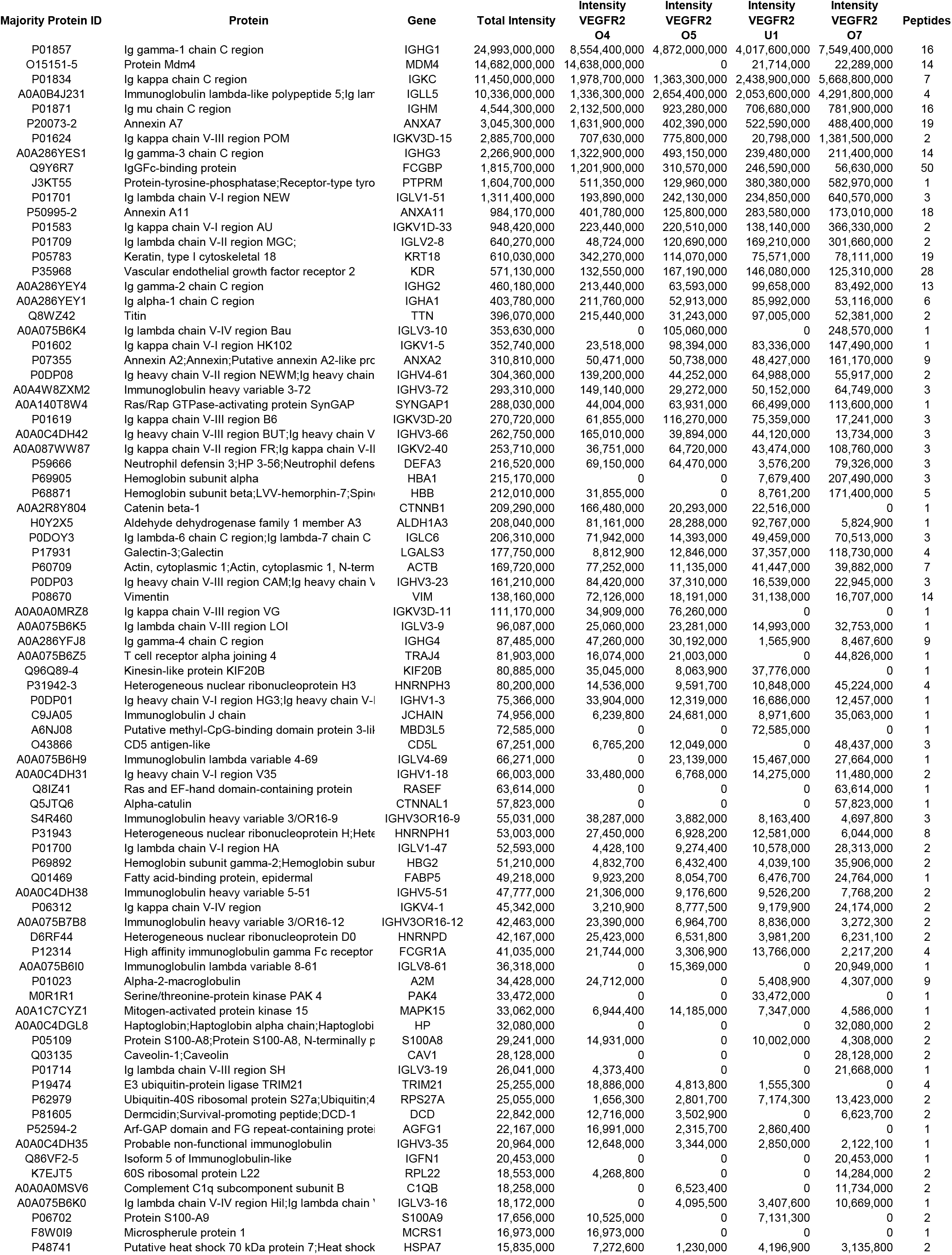
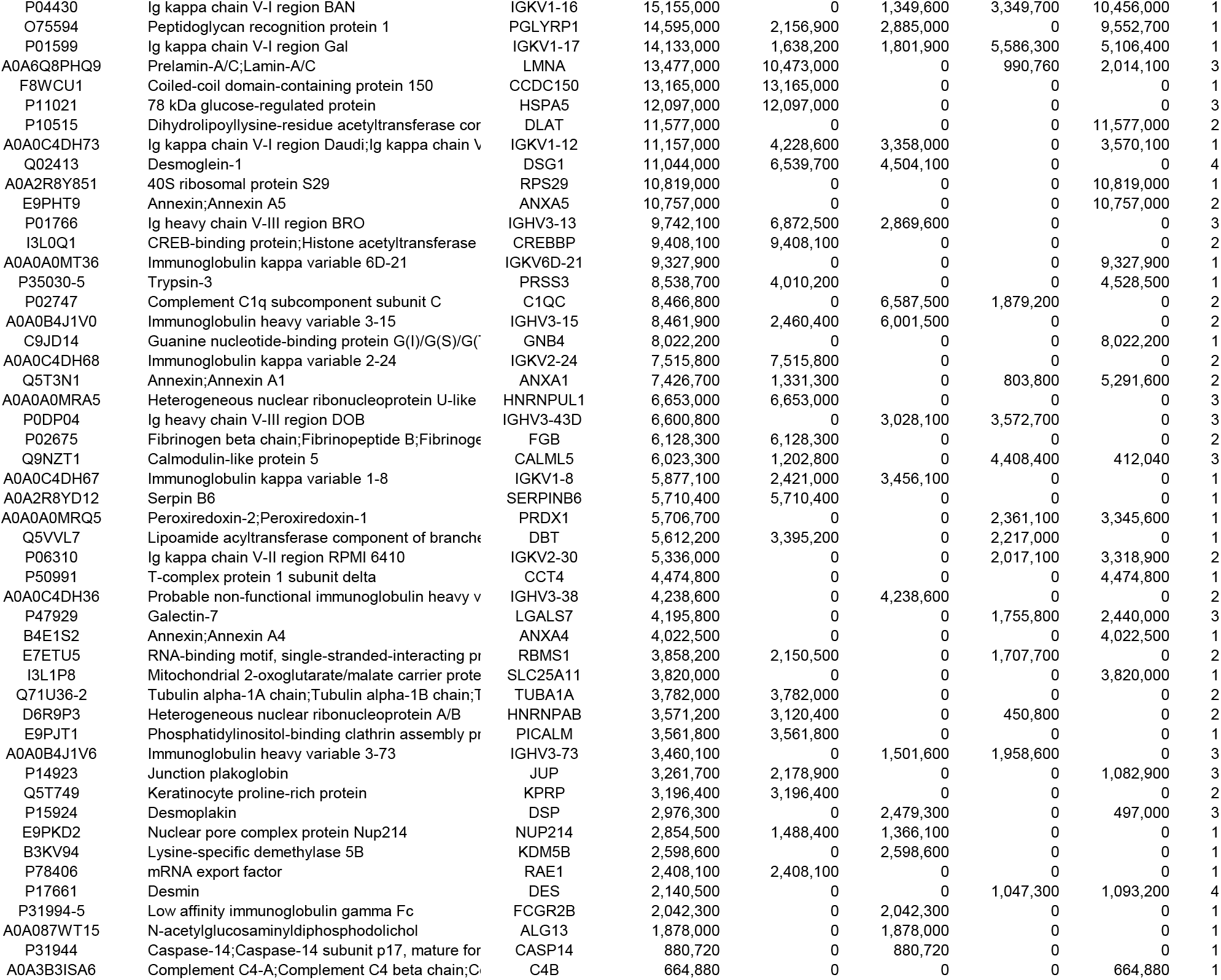
Proteins identified in VEGFR2 immunoprecipitations. Placental extracts O4, O5, U1, and O7 were immunoprecipitated in one experiment.

### Select proteins that co-immunoprecipitated with VEGFR2 (Table 2)

VEGFR2 was retained on the beads by an Ab raised to the cytoplasmic tail of VEGFR2 exposing extracted proteins to its extracellular, VEGF-binding, domain (see Methods). Therefore, most VEGFR2 partners must have interacted, even after VEGFR2 internalization, with its extracellular rather than its intracellular tyrosine kinase domain, and in binary associations while fewer would bind via some intermediate protein partners. In principle, VEGFR2, MDMX, and PICALM isoforms post-translationally modified at the target sequence of the immunoprecipitating antibodies, are unlikely to be retained on the beads. Our experimental design provided only an estimate of peptide levels in each placental sample. Very basic proteins, such as VEGF-A (pI=9.2), were not detected due to excessive trypsin digestion of these mostly intrinsically disordered proteins prior to mass spectrometric analysis. For example, more than 65% of the VEGF-A residues are expected to be disordered. Nevertheless, western blot analysis showed that VEGF-A co-immunoprecipitated in VEGFR2 IP, and that OT-R (pI=9.6) and V1aR (pI=9.5) co-immunoprecipitated in MDMX and PICALM IP. p53 and MDM2 were detected in IP eluates only by western blots. Under our protocol, proteins larger than 220 kDa were, most likely, identified by peptides from their smaller forms in the placenta.

The E3 ubiquitin ligase TRIM21 (tripartite motif-containing protein 21) co-immunoprecipitated with VEGFR2 and in MDMX and PICALM IP (Tables 2-4). TRIM21, a member of the large TRIM family, contains a zinc-binding as well as other motifs. TRIM21 is found in the cytosol and nucleus and is unique among all proteins as the highest-affinity Fc receptor in humans^26^. TRIM21 does not distinguish free from bound antibodies. Another VEGFR2 partner was PDC-E2 (*DLAT*), the E2 component of pyruvate dehydrogenase (Table 2). The association of PDC-E2 with VEGFR2 probably occurs in the nuclei^27^ and mitochondria, as discussed later, and is shown in the vasculature of the villi (video). Other VEGFR2 partners were complement components (Table 2) revealing complement activation known to occur in placental dysfunctions^28^. Immunoglobulin heavy constant alpha 1 (*IGHA1*), an autoantibody antigen and signature protein of plasma cells^29^, was among the large amounts of immunoglobulins detected that are carried by placental endothelial cells^30^ and maternal blood cells. A smaller than 220 kDa form of the giant protein titin was also detected in VEGFR2 and PICALM IP (Tables 2, 4).

### Select proteins that co-immunoprecipitated with MDMX and PICALM (Tables 3, 4)

VEGFR2 was not detected in MDMX and PICALM IP, probably because of the limited binding capacity of the MDMX and PICALM antibody charged on the beads. Among the proteins that co-immunoprecipitated with MDMX were PICALM, annexins, arginase-1, RNA-binding protein HNPNPA2B1^31^ and others that immunoprecipitated also in VEGFR2 and PICALM IP (Tables 2, 4). Protein-glutamine gamma-glutamyltransferase 2 *(TGM2)*, which co-immunoprecipitated with MDMX and PICALM (Tables 3, 4), catalyzes protein cross-linking, is considered a bridge between inflammation and hypertension, and is upregulated in preeclampsia^32^. We indicated in Table 3 the few proteins common in VEGFR2, MDMX and PICALM immunoprecipitations, and many more that co-immunoprecipitated only with MDMX, likely members of the HC proteome. After an initial statistical analysis associated MDMX with the mode of delivery, we were prompted to study the OT-R, which is activated in the myometrium causing uterine contractions^22^, and its partner V1aR, after validating two commercial antibodies (Figure 10).

**Table 3.**
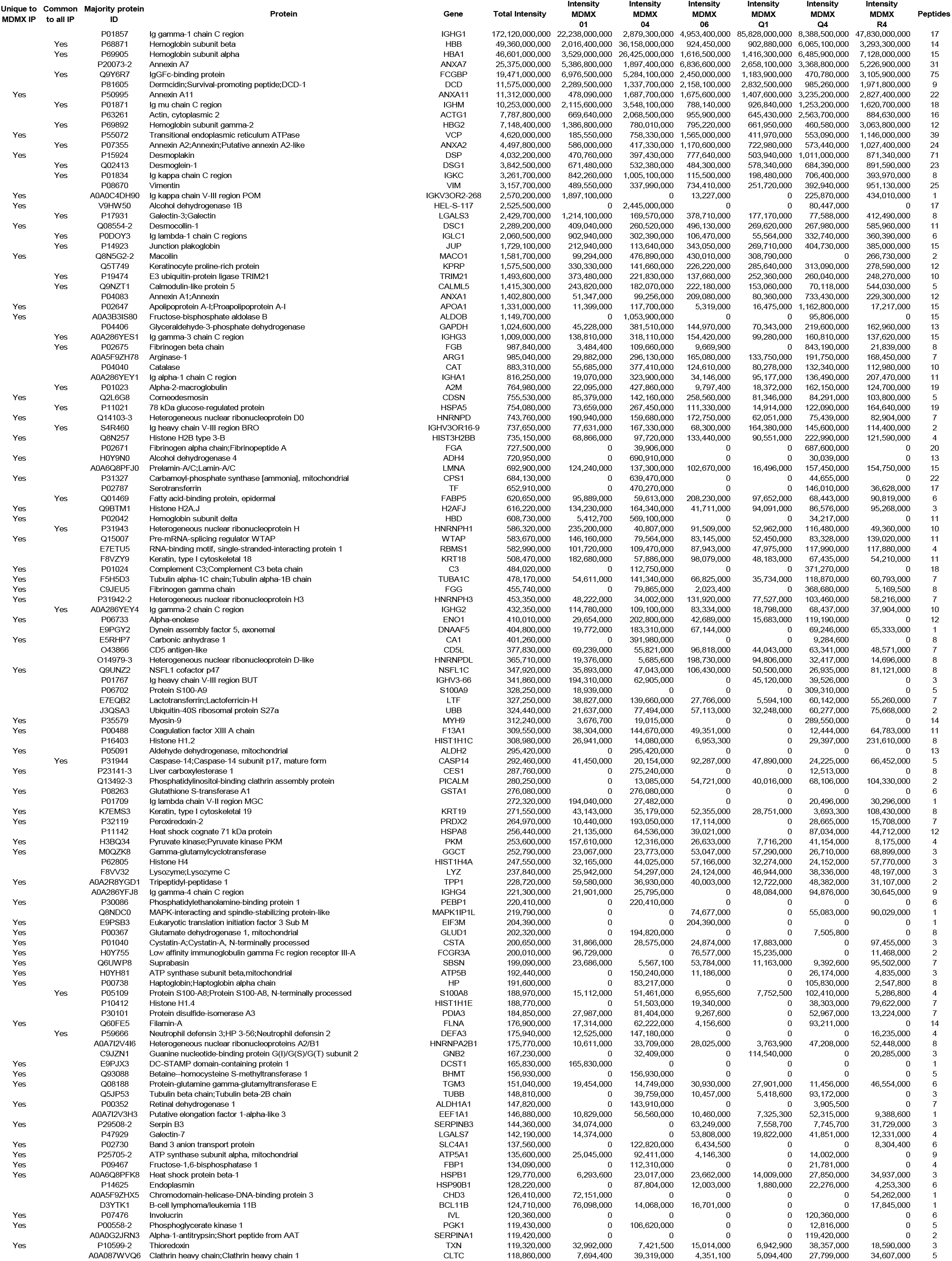
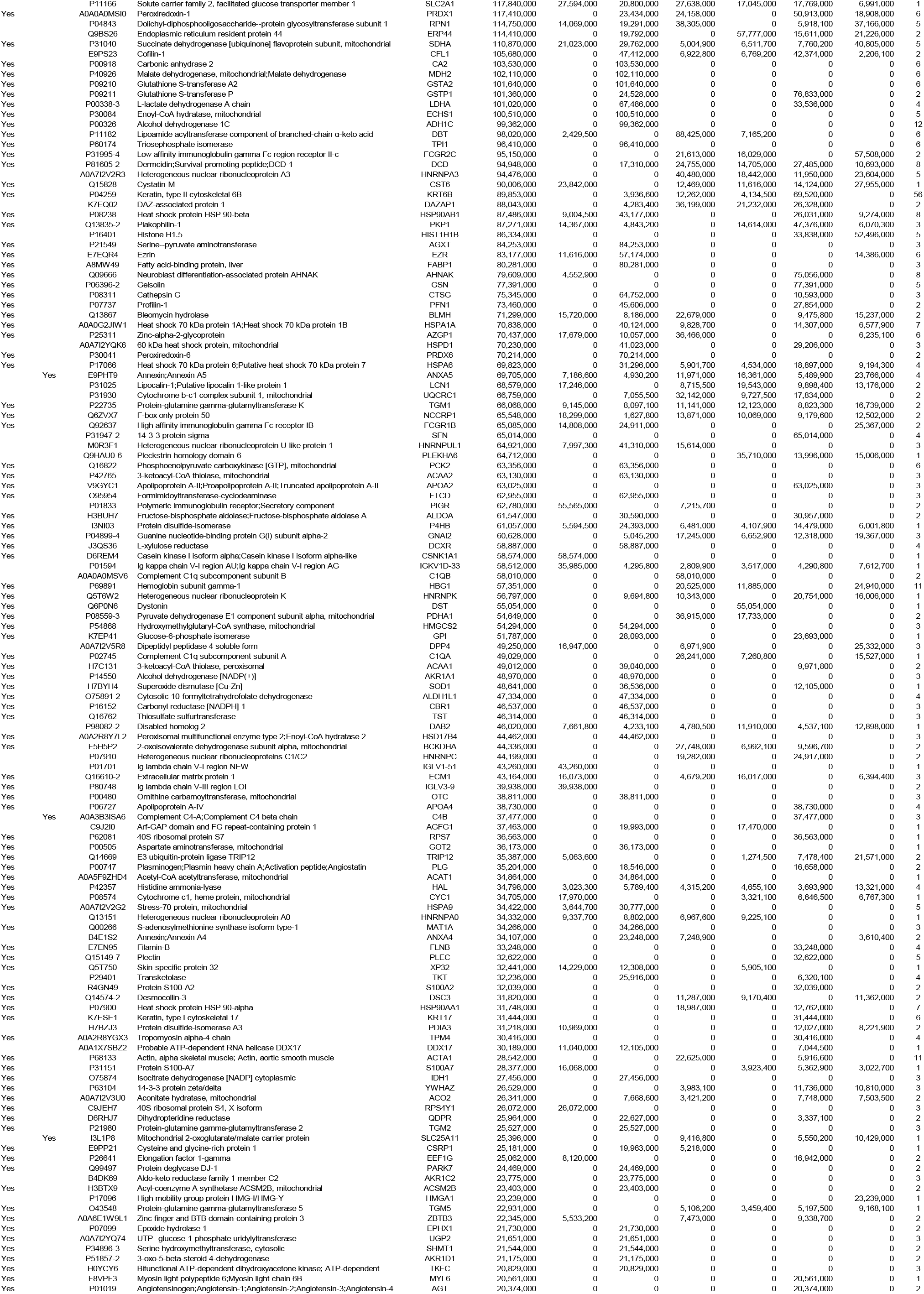
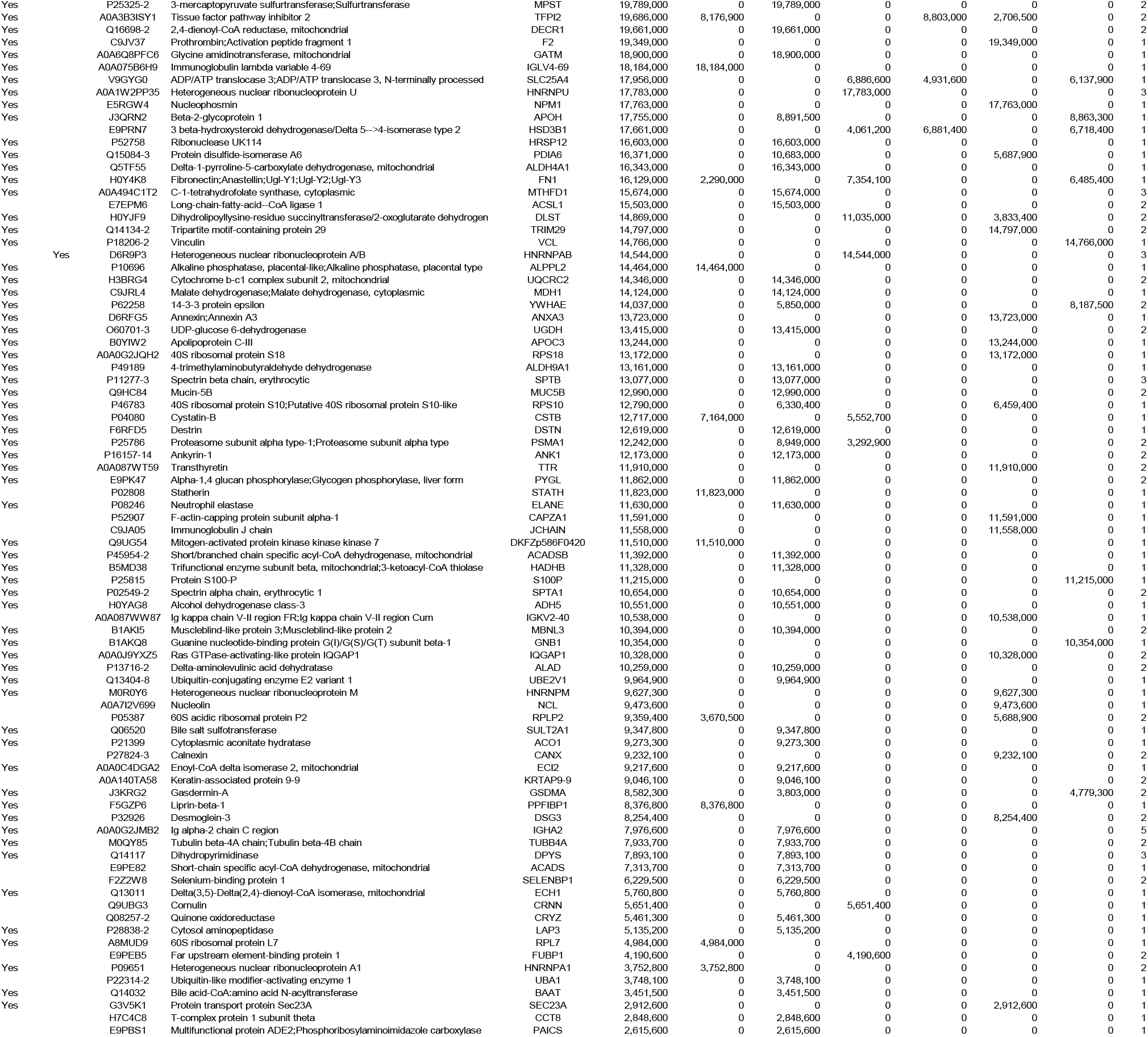
Proteins identified in MDMX immunoprecipitations (IP). Protein extracts O1, O4, O6, Q1, Q4, and R4 were immunoprecipitated in one experiment.

**Table 4.**
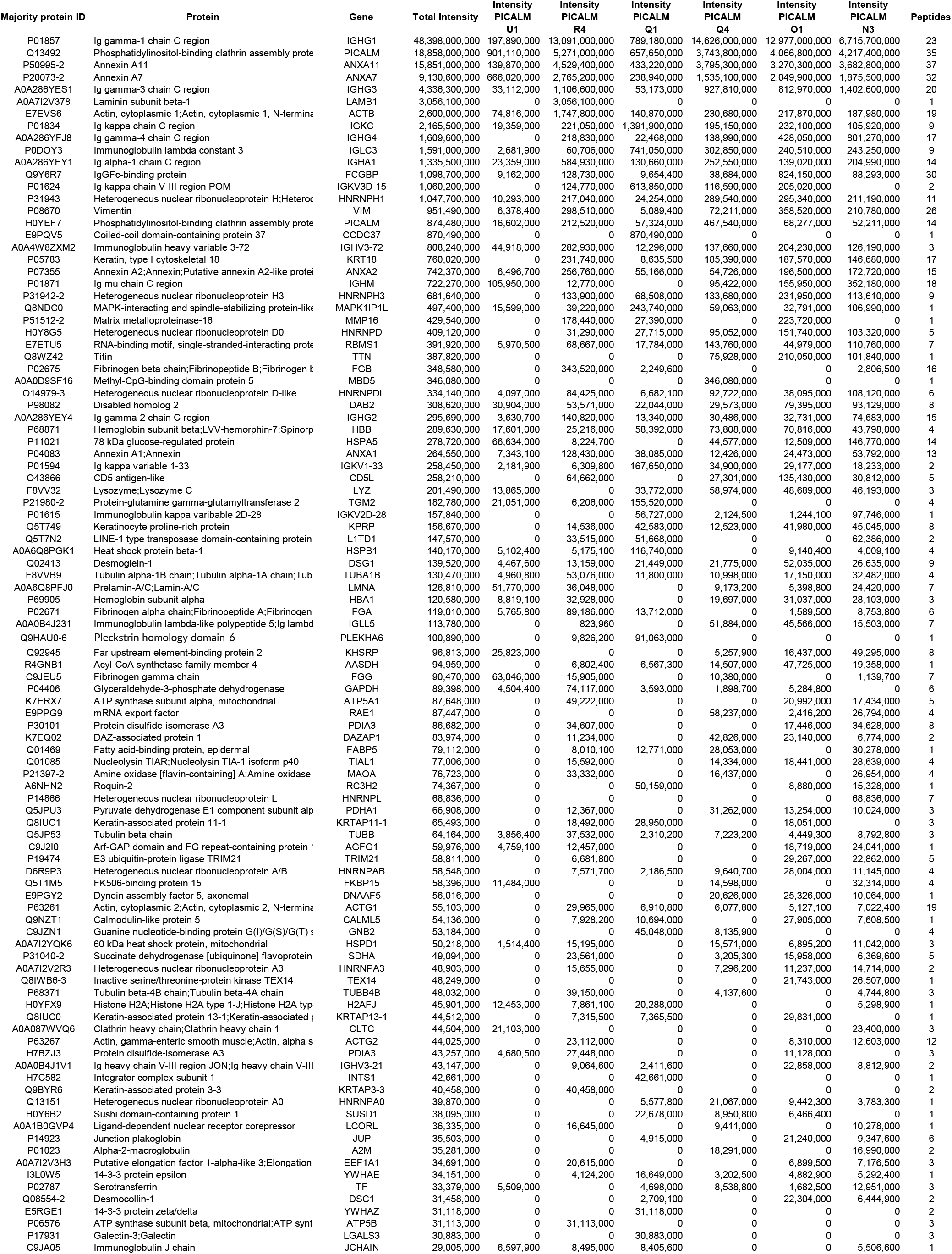
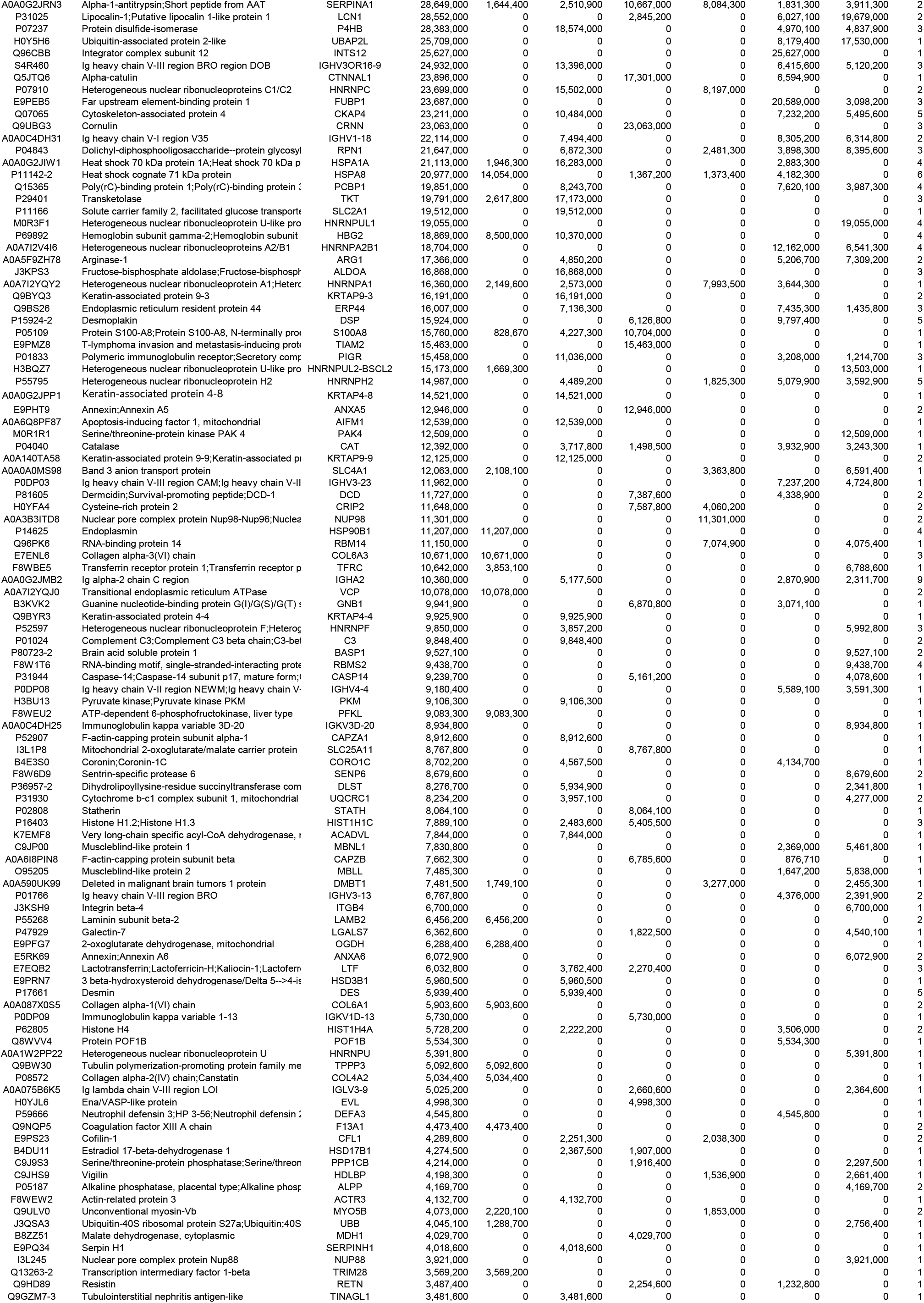
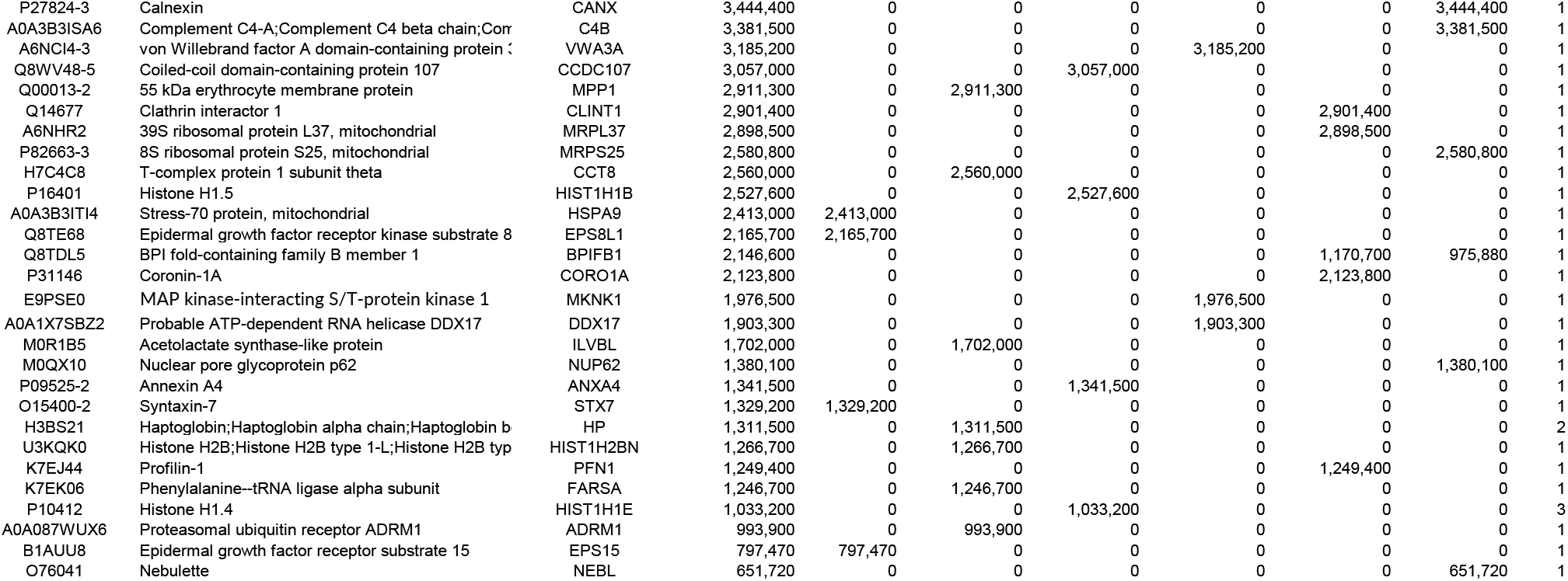
Proteins identified in PICALM immunoprecipitations. Placental extracts U1, R4, Q1, Q4, O1, and N3 were immunoprecipitated in one experiment.

### Immunohistochemistry and whole mount immunofluorescence

VEGFR2 strongly stained the endothelium of the villous capillaries (Fig. 2A). TRIM21 staining is seen in the cytoplasm of villous trophoblasts and stronger staining in intervillous maternal leukocytes (Fig. 2B, arrows). MDMX is predominantly expressed (Fig. 2, C and D) on Hofbauer cells (HC)^33–37^ that are targets of Zika and other viruses^38^. Strong MDMX staining was limited to the cytoplasm of HC, easily identified within the villous stroma, some within stromal channels (Fig. 2C). CD163^33, 39^, a marker for placental macrophages^40^, stained the cytoplasm of the HC (Fig. 2D). PICALM showed strong staining of trophoblasts and syncytiotrophoblasts (Fig. 2E) and on higher power images (Fig. 2F) positive cytoplasmic staining of the villous capillary endothelial cells was also seen.

**Fig. 2.**
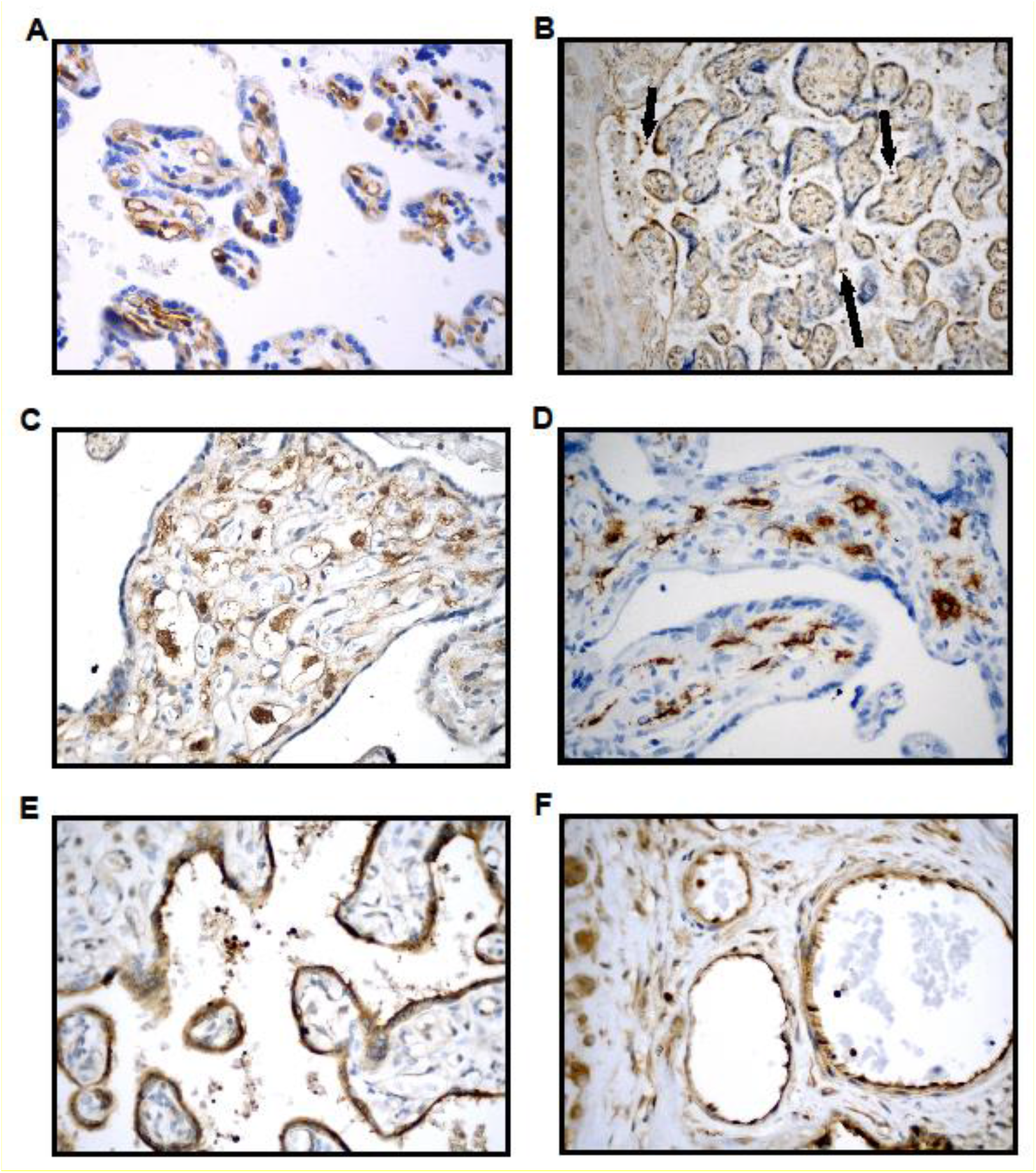
**Immunohistochemical staining of placental sections. A**. VEGFR2 immunostaining is strongly positive in villous endothelial cells (patient Q3). **B**. TRIM21 is uniformly positive in villous trophoblast (patient N4). Very strong staining is seen of maternal leukocytes (arrows). **C.** MDMX is strongly positive in the cytoplasm of HC and moderately positive in endothelial cells (patient W2). **D.** CD163 is strongly positive in the cytoplasm of the HC (patient U1). **E.** PICALM is strongly positive in the trophoblast (patient R4). **F.** PICALM positivity seen in the villous endothelial cells and fetal blood leukocytes (patient R4). Magnifications in A-F were 10x, 20x and 40x.

The co-localization of VEGFR2 (red) and PDC-E2 (green) in endothelial cells of the villous vasculature of a normotensive patient is shown on the video obtained from reconstructed stacked images of whole mount immunofluorescence. Nuclei were stained blue with DAPI.

### Immunogold electron microscopy (IGEM)

VEGFR2 was localized along segments of endoplasmic reticulum, and in mitochondria (Fig. 3). MDMX was localized diffusely within the cytoplasm of an HC and was seen clustering in the nucleus and cytosol. Clusters ran along the nuclear membrane and appeared associated with mitochondrial and endoplasmic reticulum membranes (Fig. 4). PICALM was localized to endothelial cell junctions, along endothelial cell plasma membrane, in cytoplasmic projection into the lumen and adjacent stroma and fetal blood (Fig. 5). OT-R was detected in endoplasmic reticulum and cytoplasmic “peninsulas” of endothelial cells extending into the lumen and on clusters (Fig. 6). V1aR was localized to endothelial cell membrane, nucleus, and stroma. V1aR was also seen on a fetal RBC (Fig. 7). Details are seen in Figures 3-7.

**Fig. 3.**
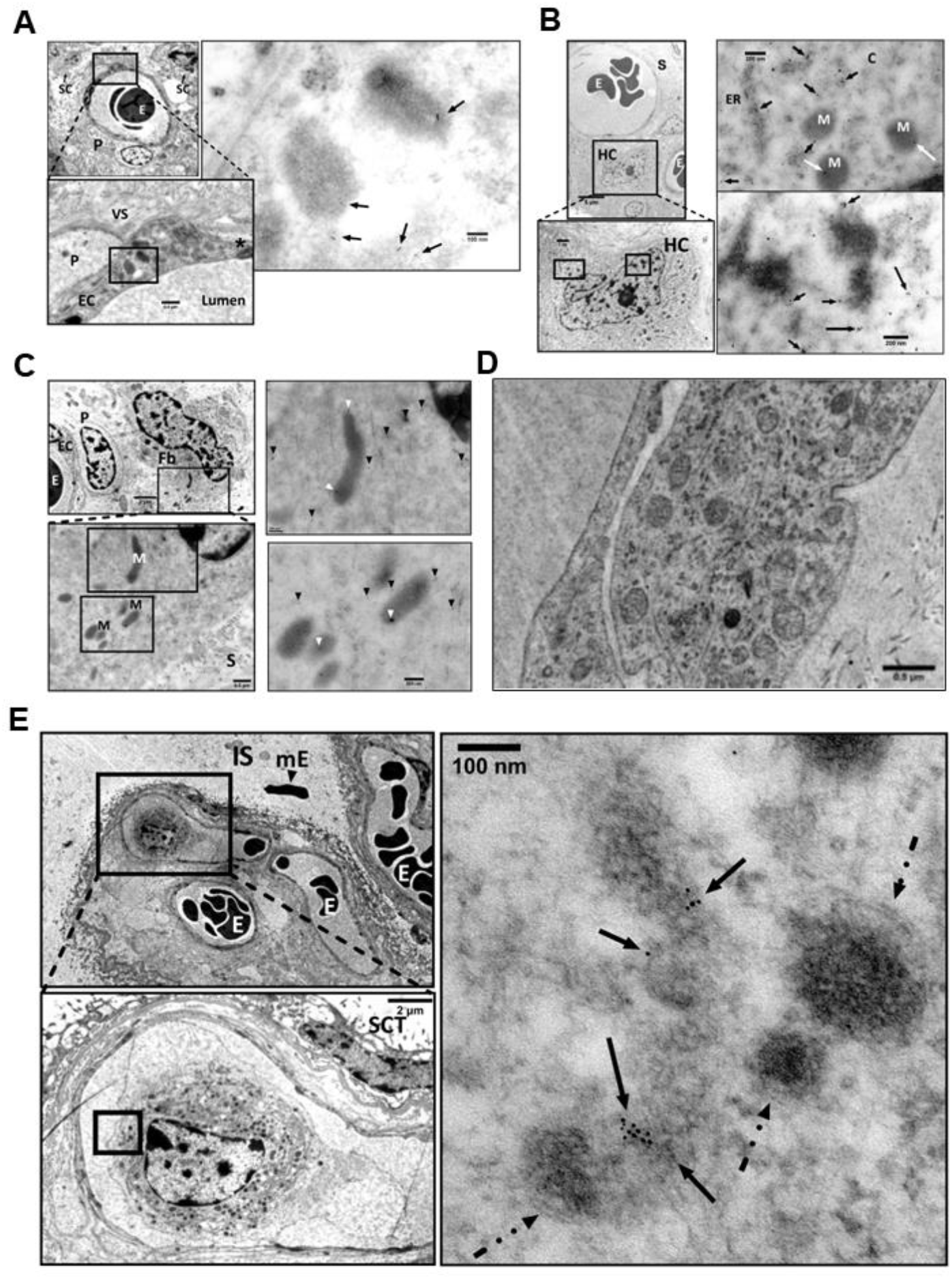
Detection of VEGFR2 by IGEM in chorionic villi of placenta Ho-73 (preeclamptic). A. Top left micrograph shows a villous capillary. P: pericyte, E: RBC, SC: stromal cells. Bottom image details area outlined on the top micrograph and shows endothelial cell (EC) cytoplasm, pericyte (P), tight junction (*), collagen fibers in villous stroma (VS), and capillary lumen (L). Image at right: Enlargement of area outlined on the bottom left shows VEGFR2 in mitochondria (arrows). **B.** Micrograph on top left shows villous capillary, stroma (S), luminal RBC (E), and Hofbauer cell (HC). Enlargement of rectangle at bottom shows prominent nucleolus, mitochondria, and endoplasmic reticulum of HC. Micrographs on the right show enlargement of areas outlined on the bottom left micrograph that detail clusters of VEGFR2 (arrows) in the cytosol (C), along segments of endoplasmic reticulum (ER), and mitochondria (M). Enlargement of image at bottom right shows diffuse localization of VEGFR2 clusters throughout the nucleoplasm (arrows). **C.** Top left micrograph: Villus stroma depicting a fibroblast (Fb), and a pericyte (P) associated with fetal capillary in partial profile, RBC (E), and EC cytoplasm. Bottom left is enlargement of area outlined on the top graph and shows fibroblast cytosol, mitochondria (M), and villous stroma (S). To the right, enlargement of two areas show VEGFR2 in cytoplasm and in partial profiles of mitochondrial matrix (white arrow heads). **D.** A TEM micrograph after osmication provided a better outline of mitochondria in an EC, compared to **A, B** and **C** panels**. E.** Fetal capillaries with luminal RBC (E) are shown in the top left image, IS: intervillous space; mE: maternal RBC. Bottom: enlargement of outlined area shows a fetal macrophage in greater detail; a syncytiotrophoblast (SCT) is also present. Right micrograph: enlargement of area in bottom left micrograph, points to VEGFR2 labelling in an incomplete profile of a mitochondrion. Dashed arrows outline the outer double membrane of the labeled and unlabeled mitochondria. The gold particle diameter is 6 nm.

**Fig. 4.**
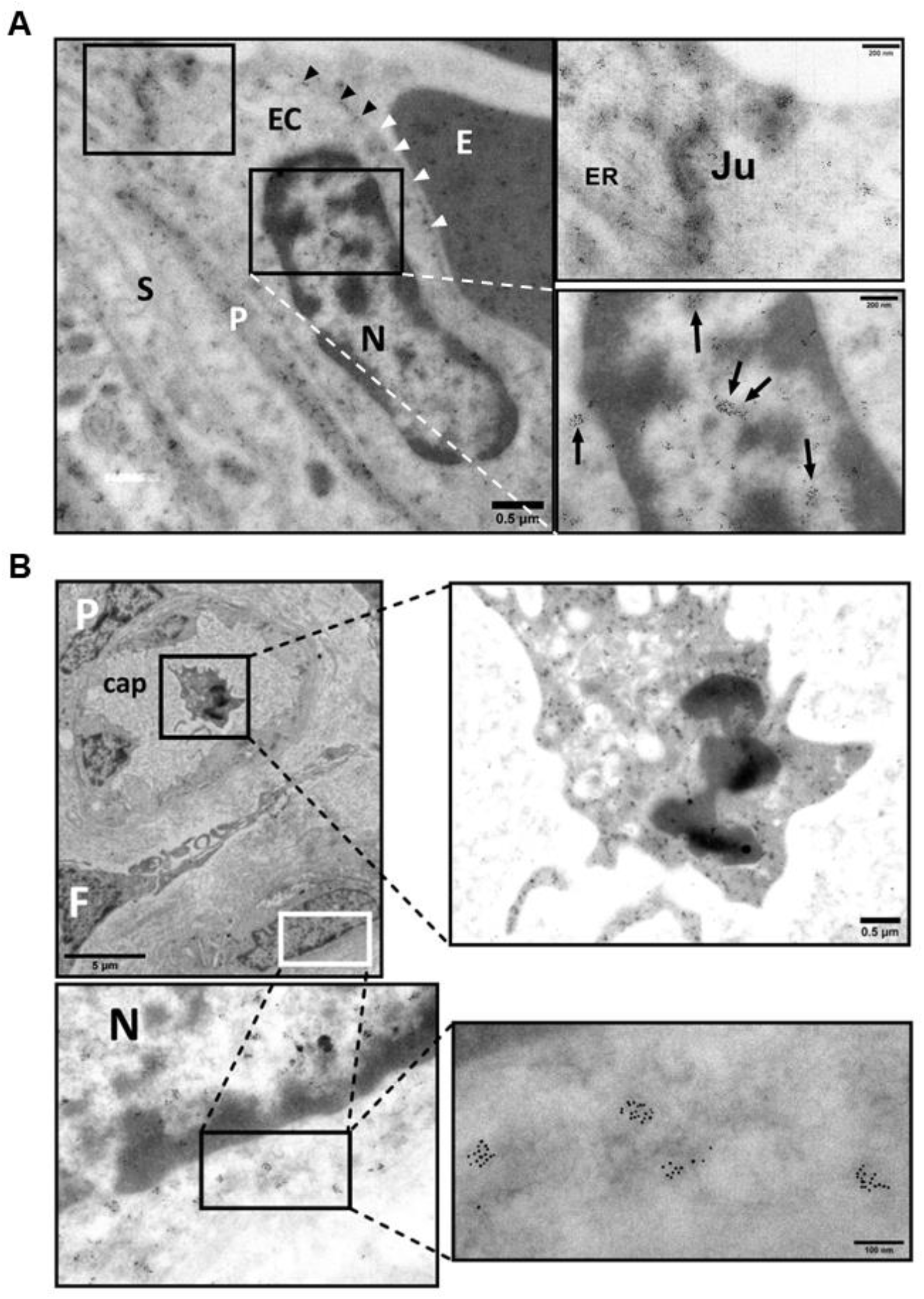
Detection of MDMX by IGEM in chorionic villi of placentas Ho-71 (normotensive) and Ho-73 (preeclamptic). A. Left micrograph: Nucleus (N) of endothelial cell (EC) from Ho-73. Arrow heads point to small MDMX clusters, parallel to the inner aspect of the EC plasma membrane. Area outlined, top left: EC junction; P: pericyte process; E: RBC in capillary lumen, S: stroma. This area is enlarged on the top right graph and shows MDMX clusters along the junction (Ju), and endoplasmic reticulum (ER). Bottom right: enlargement of outlined area shows diffuse pattern of MDMX clusters throughout the nucleoplasm. **B.** Top left micrograph shows an outline of apoptotic macrophage in the lumen of fetal capillary (cap) from Ho-71. P: pericyte, F: partial profile of fibroblast (F) and HC partially marked by a white outline. Enlargement on the top right image shows diffuse MDMX localization in the vacuolated cytoplasm of luminal macrophage. Image at bottom left is enlargement of area outlined in white and shows MDMX clustering in the nucleus (N) and cytosol of the HC. Clusters, averaging 50-100 nm in diameter, run along the nuclear membrane and appear to be associated with mitochondrial and endoplasmic reticulum membranes. Bottom right: enlarged area shows MDMX clusters of approximately 100 nm. The gold particle diameter is 6 nm.

**Fig. 5.**
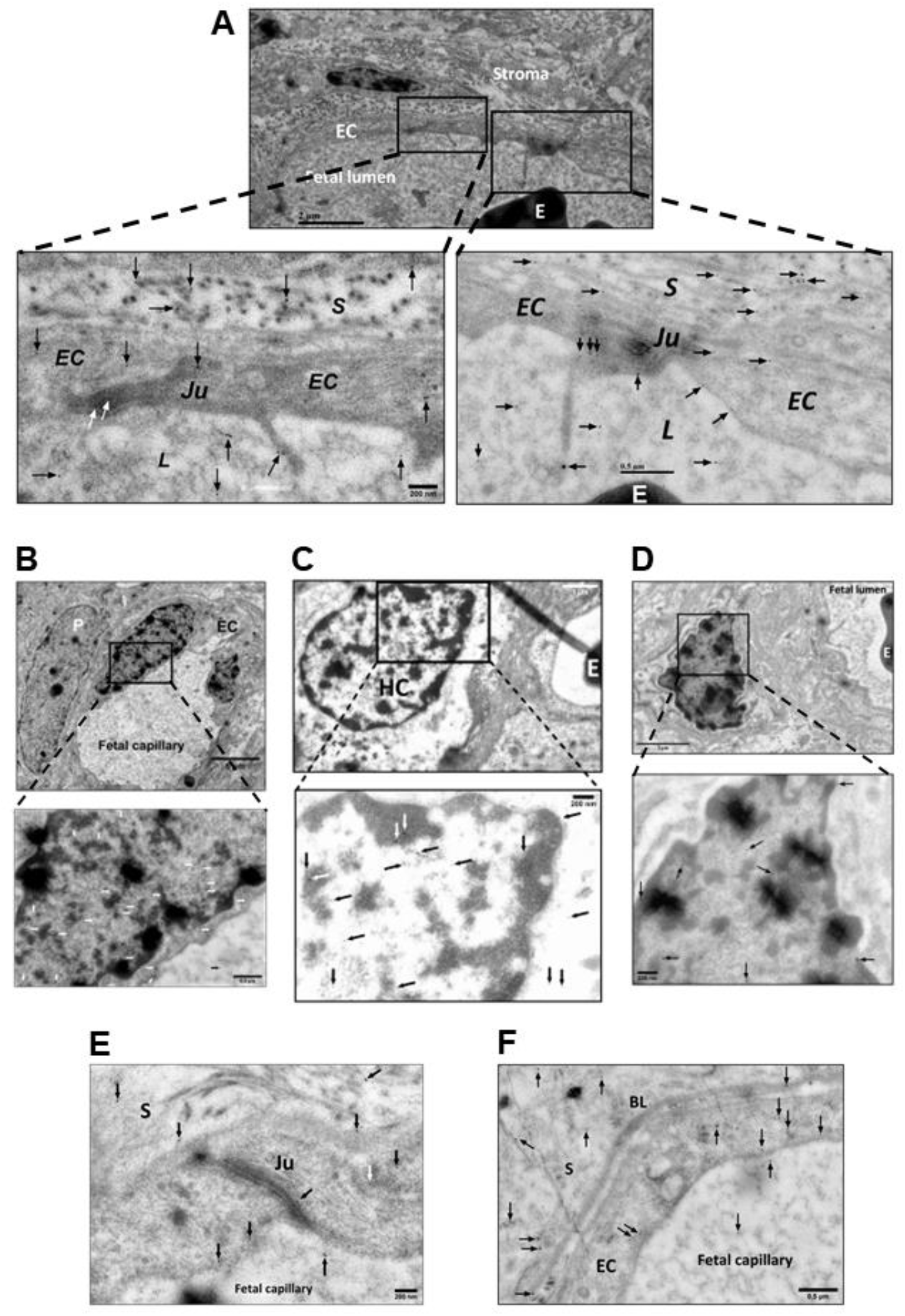
Detection of PICALM in chorionic villi of placentas Ho-71 (normotensive) and Ho-73 (preeclamptic). A. Top IGEM micrograph: partial profile of villus capillary from Ho-71. EC: endothelial cells; E: RBC; stroma. Bottom images: enlargement of two selected areas of endothelium show PICALM localization (arrows) at EC junctions (Ju), along EC plasma membrane, in cytoplasmic projections into lumen (L) and adjacent stroma (S). **B.** Top micrograph: the nuclei of endothelium of a villus capillary from Ho-71, and the adjacent pericyte (P) are patent. Bottom image: enlargement of nucleus, shows PICALM in nucleoplasm near chromatin, on EC cytoplasmic membrane (white arrows) and in luminal space (black arrow). **C.** Top micrograph: HC in stroma of Ho-73. E: RBC in capillary. At bottom: enlargement shows PICALM in HC nucleus (arrows). **D.** Top micrograph: HC cell in the stroma of placenta Ho-71. Bottom image: enlargement shows PICALM in nucleoplasm in association with chromatin. **E.** PICALM is shown (arrows) at the junction (Ju), and plasma membrane of EC and in adjacent stroma (S) of fetal capillary from Ho-73. **F.** PICALM is shown (arrows) in cytoplasm and plasma membrane of EC of capillary from Ho-71, and in basal lamina (BL), stroma (S) and capillary lumen. The gold particle diameter is 10 nm.

**Fig. 6.**
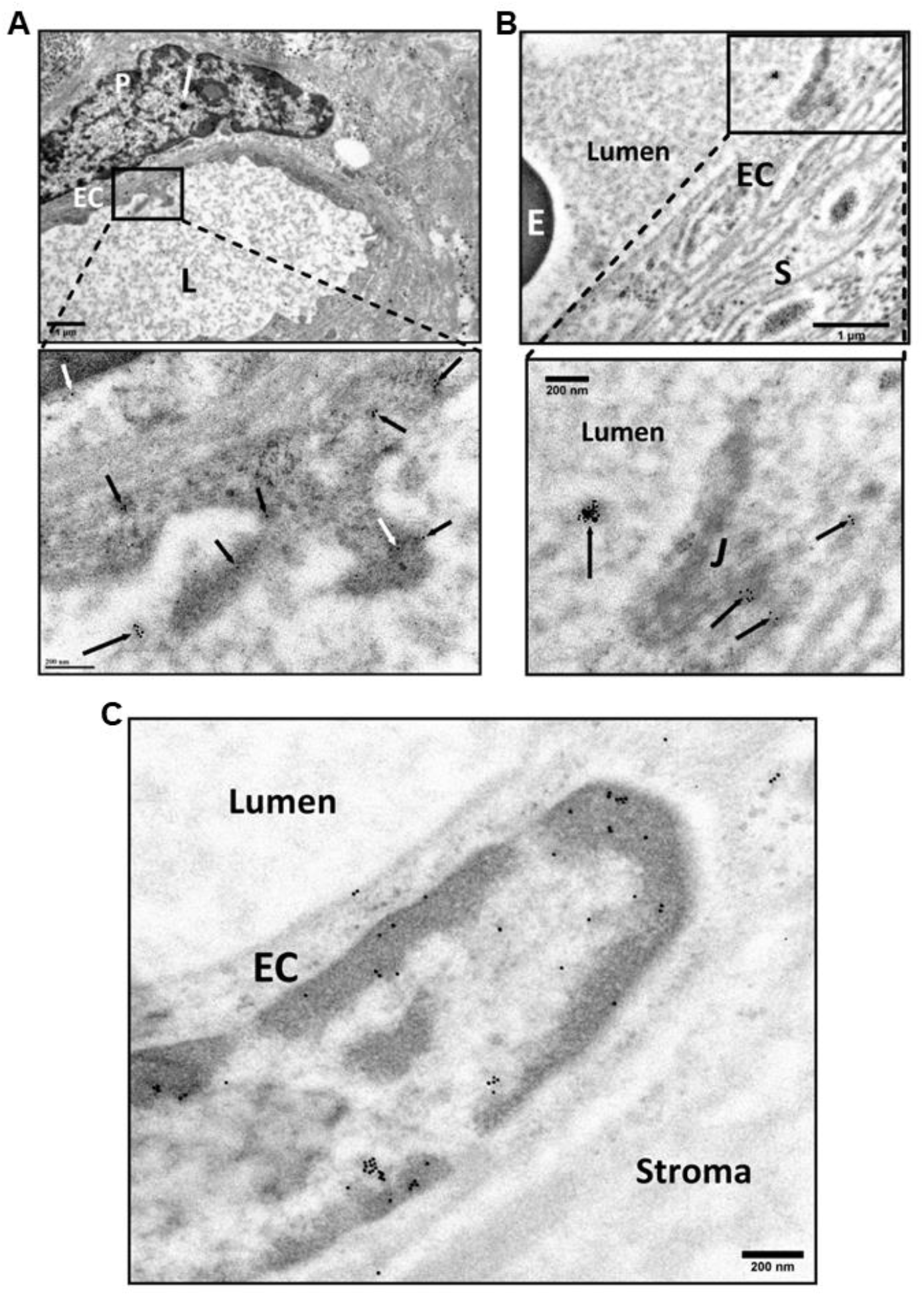
Detection of Oxytocin Receptor (OT-R) in chorionic villi of placentas of normotensive Ho-71, and preeclamptic Ho-73 patients. A. Top IGEM micrograph: Villus capillary from Ho-71. P: pericyte adjacent to the endothelial cell (EC); L: lumen. White arrow shows cluster of PICALM on P. Bottom: enlargement shows OT-R in endoplasmic reticulum (ER) and cytoplasm projections of EC into the lumen. (arrows). **B.** Top micrograph: Partial view of villus capillary from Ho73, E: RBC; S: stroma. Bottom graph shows EC junction (J) and OT-R clusters on J, EC cytoplasm, and lumen (arrow). **C.** OT-R is seen in the nucleus of an EC from Ho-73.The gold particle diameter is 10 nm.

**Fig. 7.**
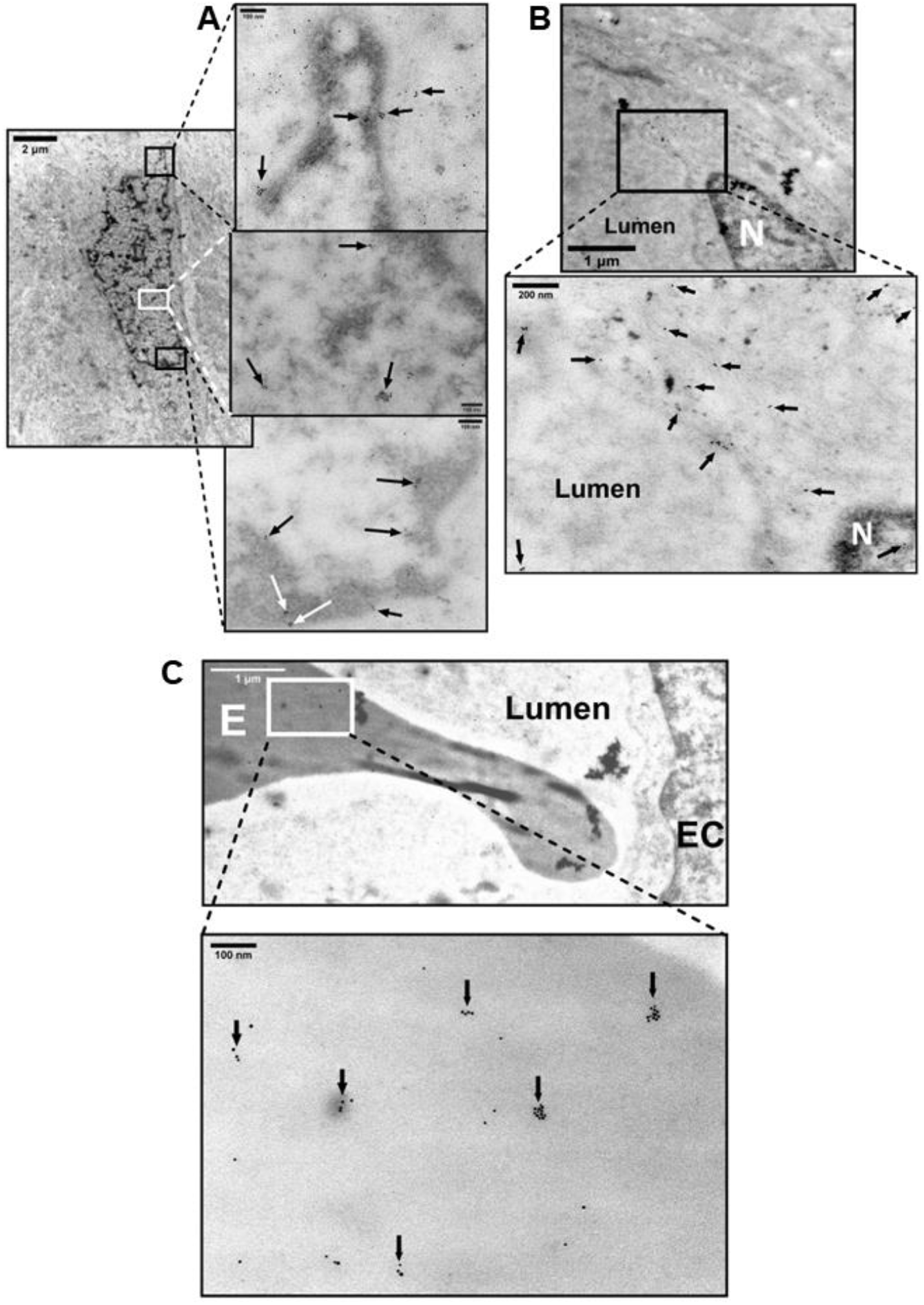
Detection of Vasopressin Receptor V1aR in chorionic villus of placenta Ho- 72 (diabetic). A. Composite IGEM image shows nucleus of HC in stroma on the left and enlargements of three demarcated regions of its nucleus on the right. V1aR clusters, 20- 100 nm, in the nucleus are indicated by arrows. **B.** Top micrograph shows EC aspect of a villus capillary. Bottom image is enlargement that shows V1aR on EC membrane, nucleus, and stroma (arrows). **C.** Top micrograph shows aspect of EC of villus capillary; an RBC is seen in the lumen. Bottom image is enlargement that shows several 20-100 nm clusters of V1aR on surface of the RBC (arrows). The gold particle diameter is 10 nm.

**Fig. 8.**
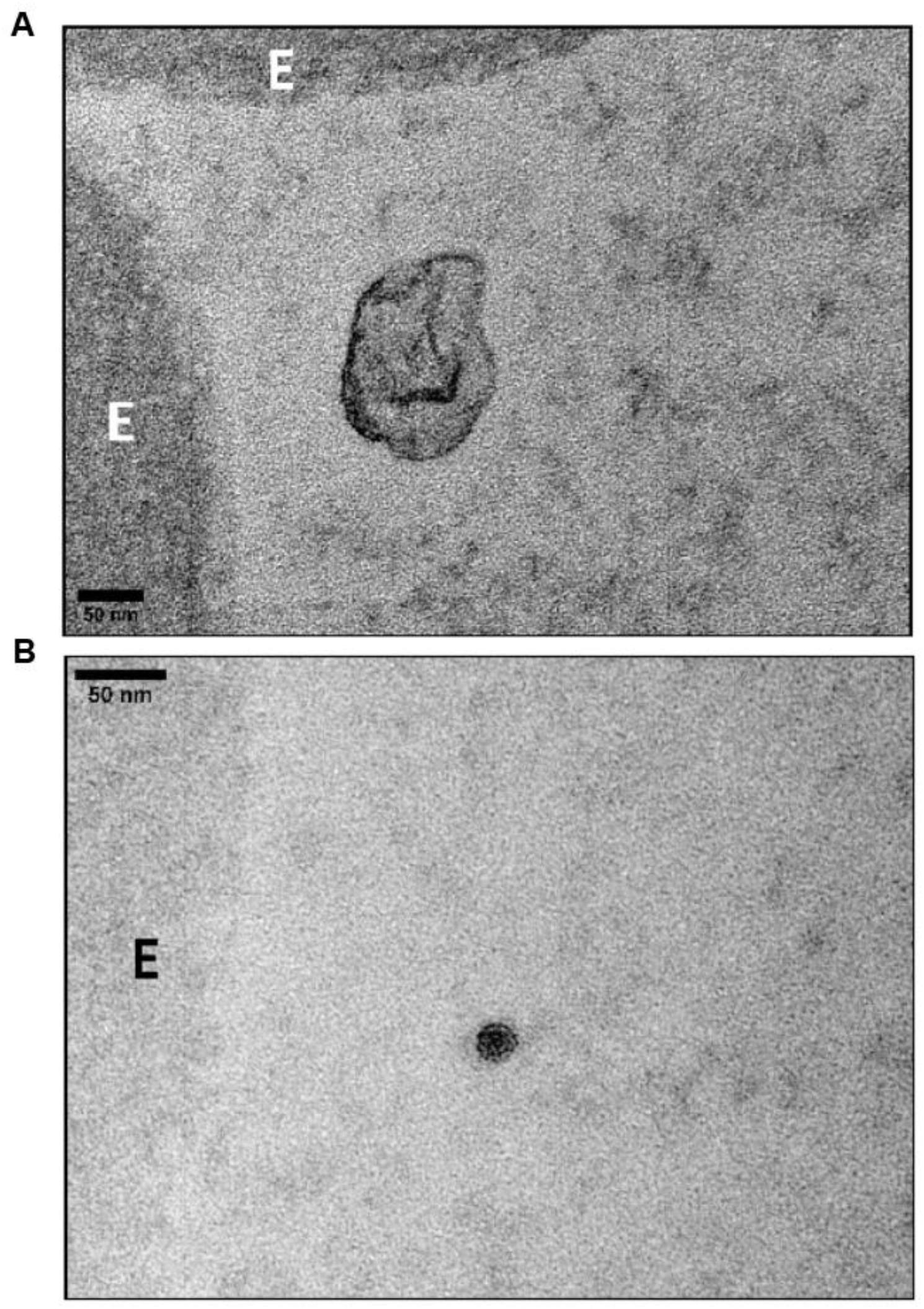
Transmission electron microscopy of osmicated chorionic villus from placenta Ho-73 (preeclamptic). A. An exosome^9^ is shown in the lumen with the characteristic lipid bilayer adjacent to RBC (E). **B.** A dense particle with a lipid bilayer, perhaps an exomere^12^, is shown in the lumen next to an RBC (E). Scale in A and B is 50 μm.

### Statistical analysis of protein levels in placental extracts

To estimate the protein levels of MDMX, PICALM, OT-R and V1aR, we analyzed by western blots 25 μg protein from each of the 44 placental extracts (Table 1). The intensity of the native-protein band, shown at the top of the representative western blots in Fig. 9, is relative to an internal control sample (Q1 or V1) taken as 100% (Table 5). Violin plots show the analysis of the mean protein levels (Fig. 9) among different clinical conditions. Precluded from the statistical analysis were VEGFR2, since its protein levels did not differ significantly among the 44 placentas analyzed in this study, and TRIM21 due to extensive degradation of its native 50-kDa form in our placental extracts.

**Fig. 9.**
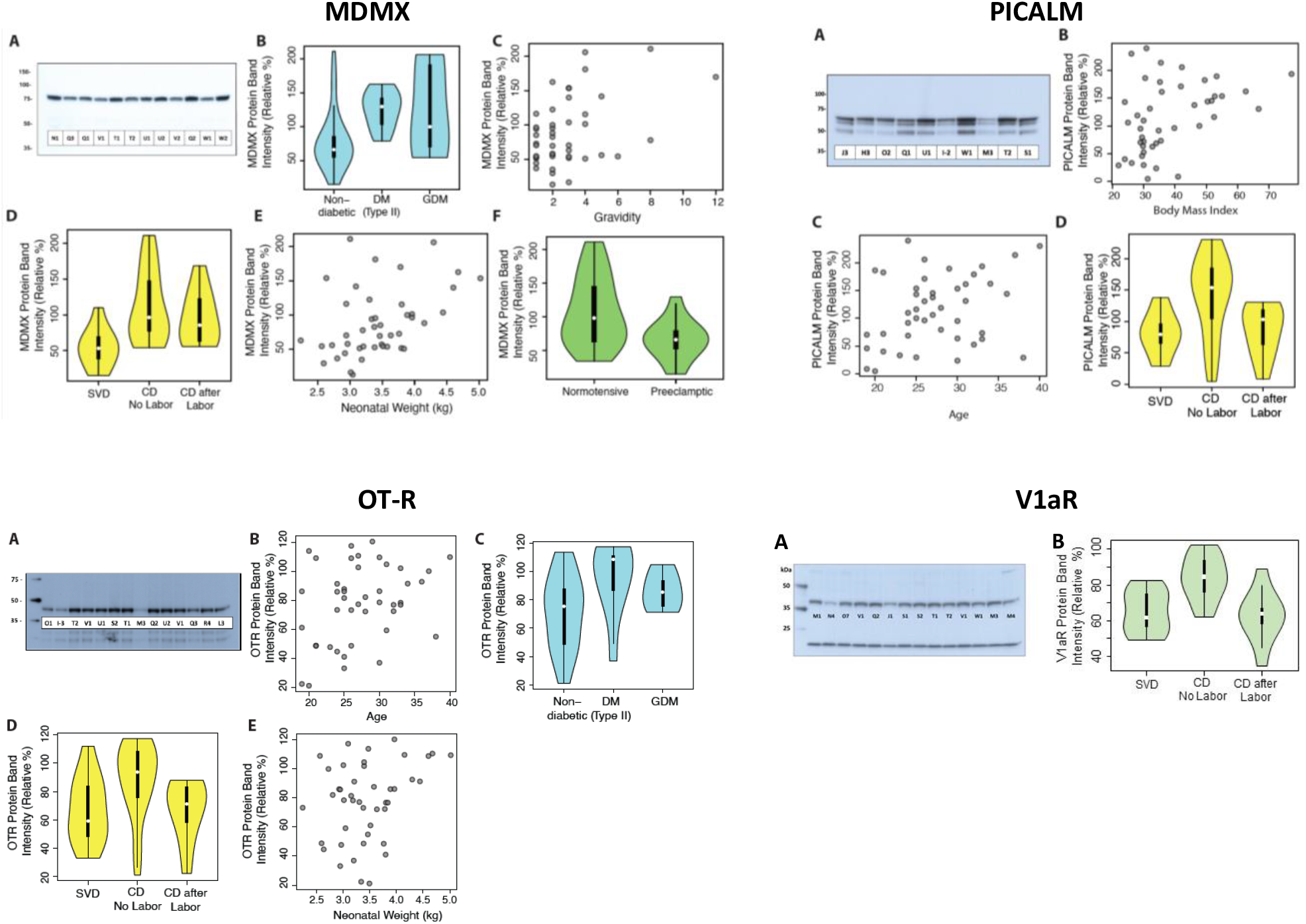
Representative western blots (A) of MDMX, PICALM, OT-R, and V1aR, and violin plots of their relative protein levels associated with diabetes, gravidity, labor, neonatal weight, preeclampsia, BMI, or maternal age, as indicated in B-E. Western blots show the molecular mass (kDa) and the placental extracts identified by a letter-number code shown in Table 1. Internal control for MDMX and PICALM was sample Q1, and sample V1 for OT-R and V1aR.

**Fig. 10.**
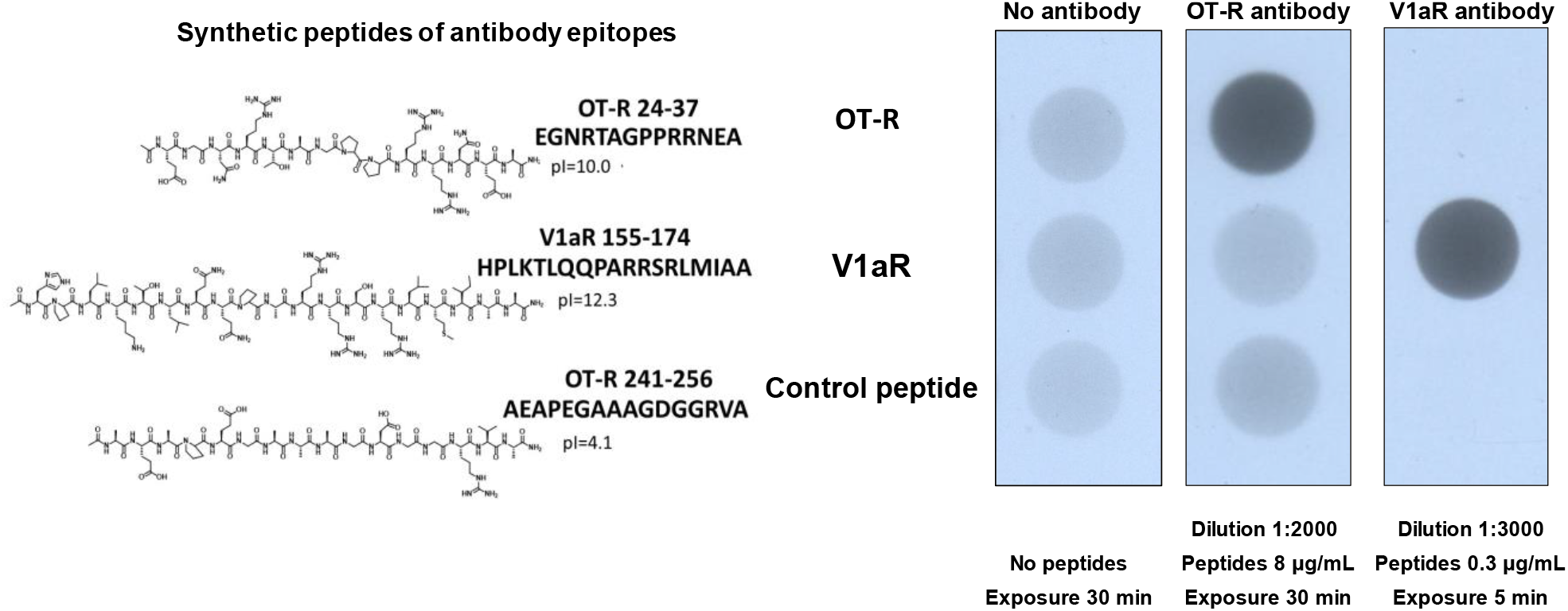
Validation of OT-R and V1aR antibodies. The synthesized peptides and their pI are shown next to the scans of the coated Nunc C-bottom strips probed with the antibodies being validated.

**Table 5.**
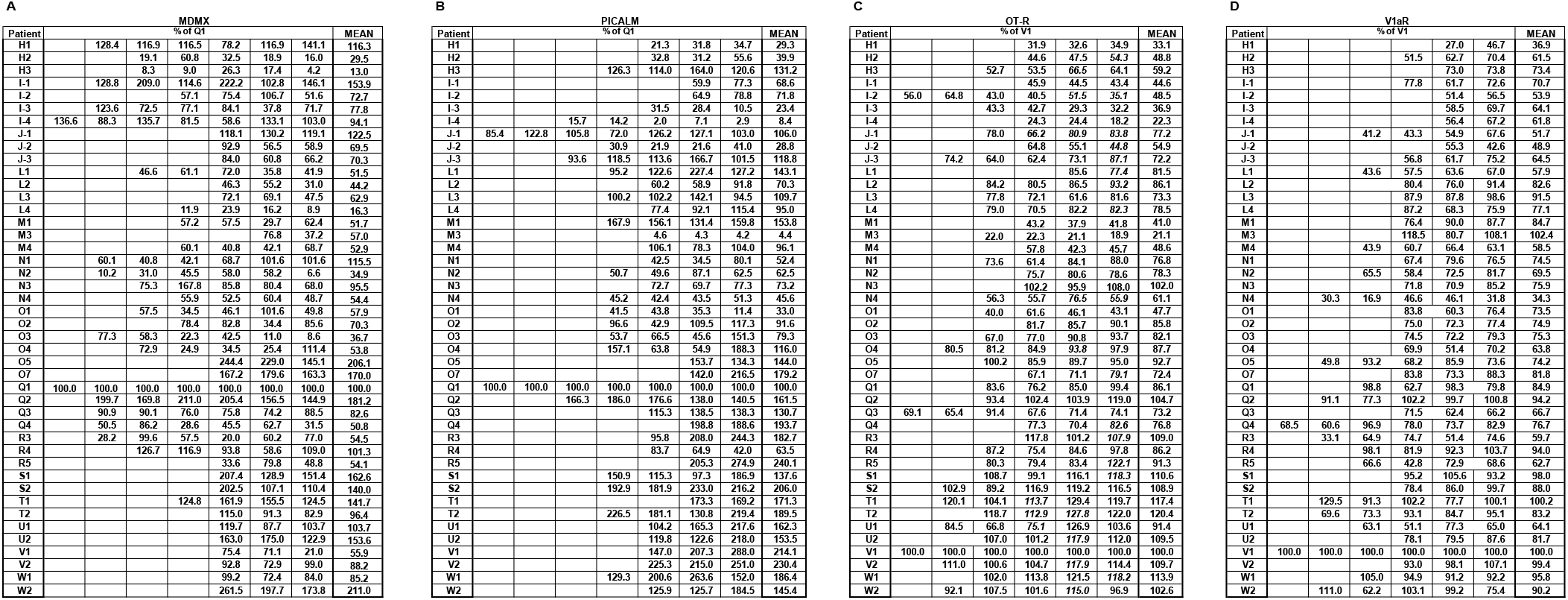
Protein levels of MDMX (A), PICALM (B), OT-R (C) and V1aR (D) in 44 placental extracts. MDMX and PICALM values are relative to extract Q1, and OT-R and V1aR values relative to sample V1. Replicate and mean values are shown.

Next, we performed univariable and multivariable analyses, shown in Tables 6-12, to test whether the mean protein levels of MDMX, PICALM, OT-R and V1aR, represented by western blot band intensity of placental extracts and relative to an internal reference sample Q1 or T1, varied as a function of maternal age, gravidity, gestational age, body mass index, race, preeclampsia, diabetic status, delivery mode, neonatal sex, and neonatal weight. Preeclamptic patients were compared to non-preeclamptic, and diabetic to non-diabetic patients.

**Table 6.**
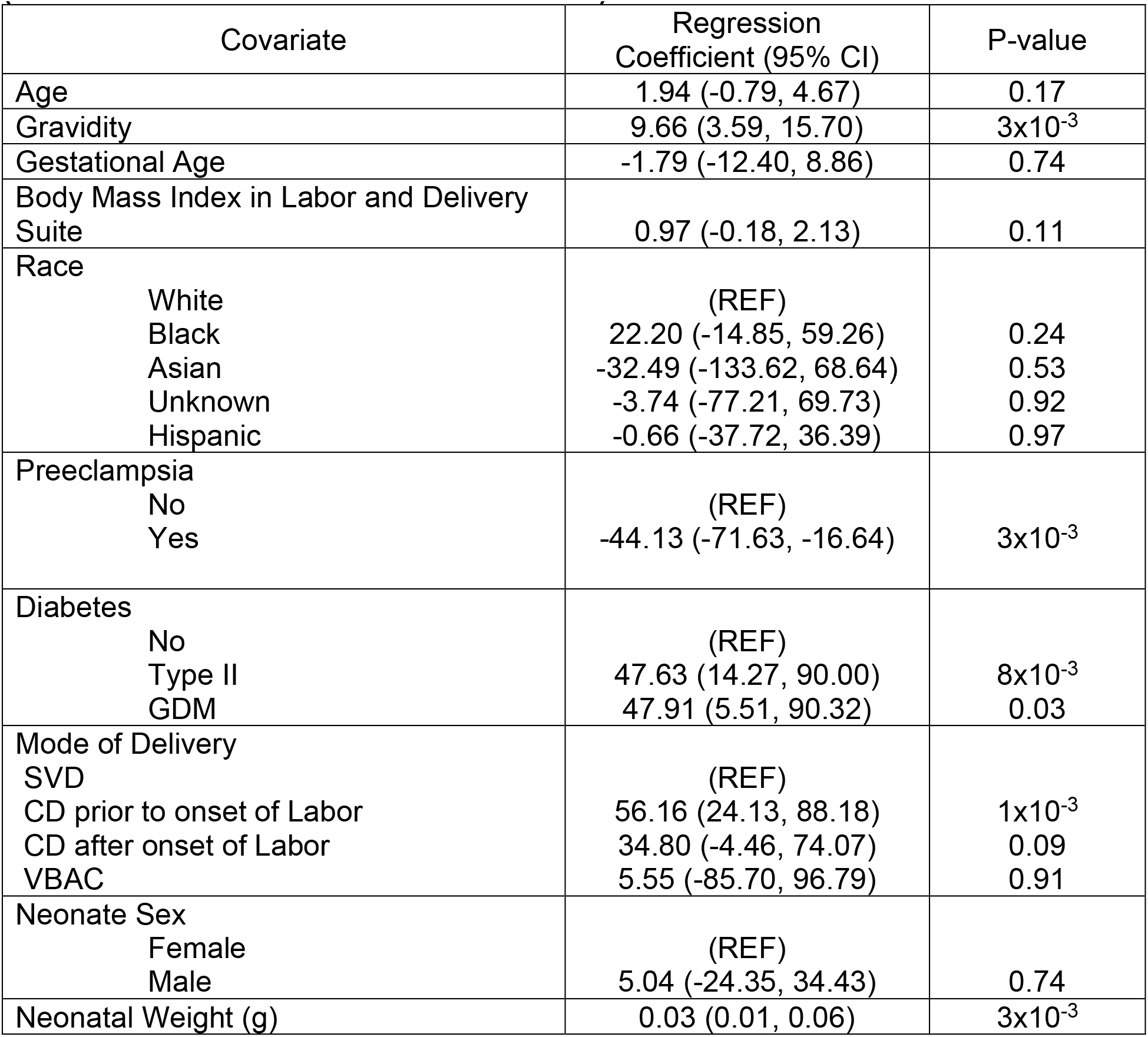
Regression coefficients from univariable analysis of MDMX protein band intensity (relative to internal reference individual Q1)

### Univariable analysis of protein levels

Protein expression units are the percent values relative to an internal control sample taken as 100% (Table 5).

MDMX protein levels were associated with diabetes with an increase of 47.63 units (95% CI 14.27, 90.00) among those with Type II diabetes and 47.91 (95% CI 5.51, 90.32) among those with gestational diabetes mellitus (GDM) compared to those without diabetes, and gravidity with an increase of 9.66 (95% CI 3.59, 15.70) with each unit increase in gravidity, neonatal weight (an increase of 0.03 (95% CI 0.01, 0.06) for each increase in grams), and preeclampsia (-44.13 (95% CI -71.63, -16.64). Regarding the mode of delivery, cesarean delivery, CD, prior to the onset of labor was associated with an increase of 56.16 (95% CI 24.13, 88.18) compared to those with SVD. See Table 6 for full results. Recently, CD163 expression in Hofbauer cells was associated with BMI, gravidity, and fetal birthweight^41^.

Increased PICALM levels were associated with maternal age with an increase of 3.68 (95% CI 0.28, 7.08) in intensity for each year increase, and body mass index with an increase of 2.06 (95% CI 0.66, 3.45) for each unit increase. Regarding the mode of delivery, CD prior to the onset of labor had an increase of 59.96 (19.24, 100.68) compared to those with SVD. See Table 7 for full results.

**Table 7.**
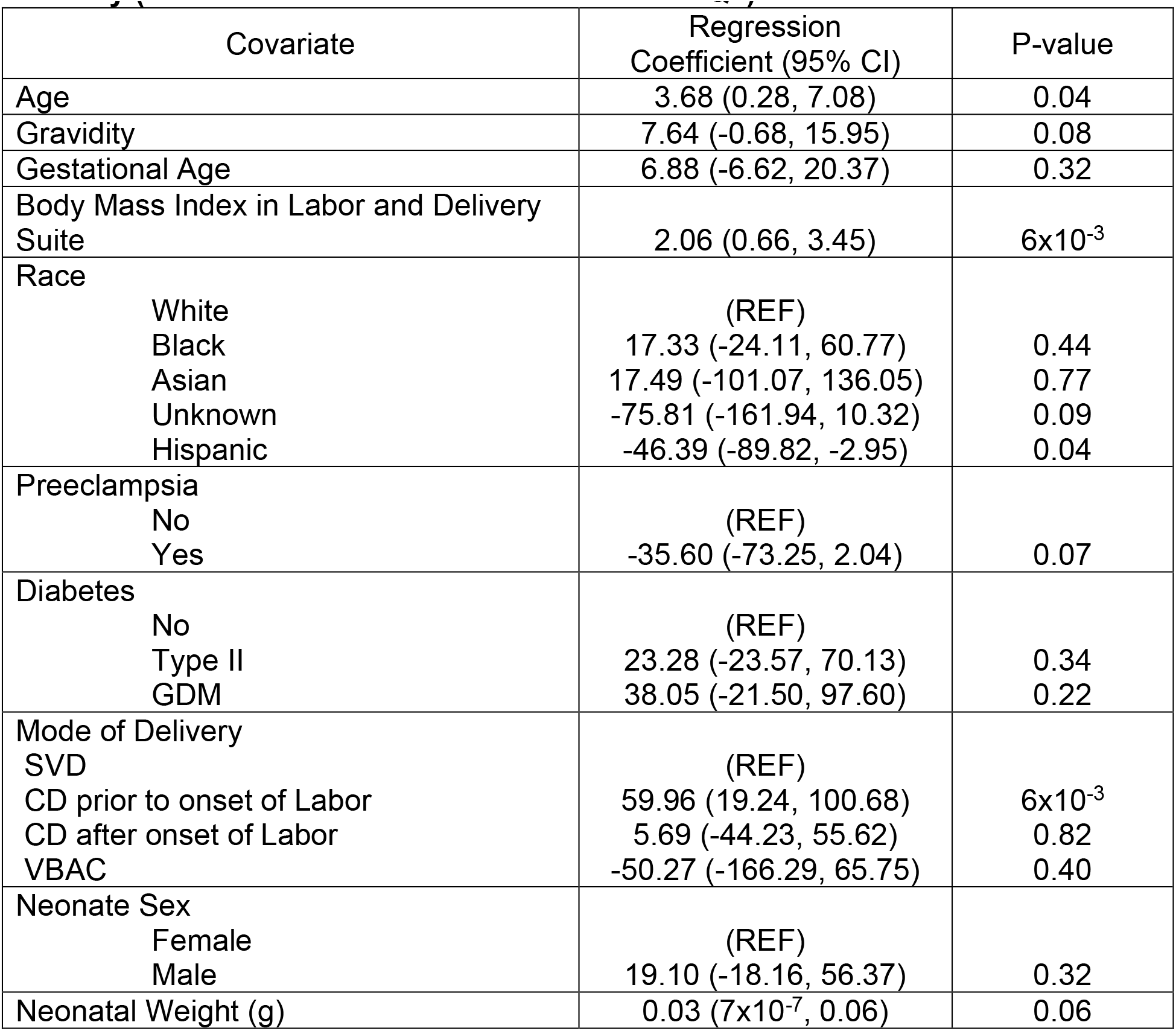
Regression coefficients from univariable analysis of PICALM protein band intensity (relative to internal reference individual Q1)

OT-R protein levels were associated with maternal age, an increase of 1.56 (95% CI 0.13, 2.99) for each year of age, and diabetes with an increase of 24.16 (95% CI 5.47, 42.85) among those with Type II diabetes compared to those without diabetes. Neonatal weight was associated with an increase of 0.01 (95% CI 8.36x10^-4^, 0.03) for each increase in grams. See Table 8 for full results.

**Table 8.**
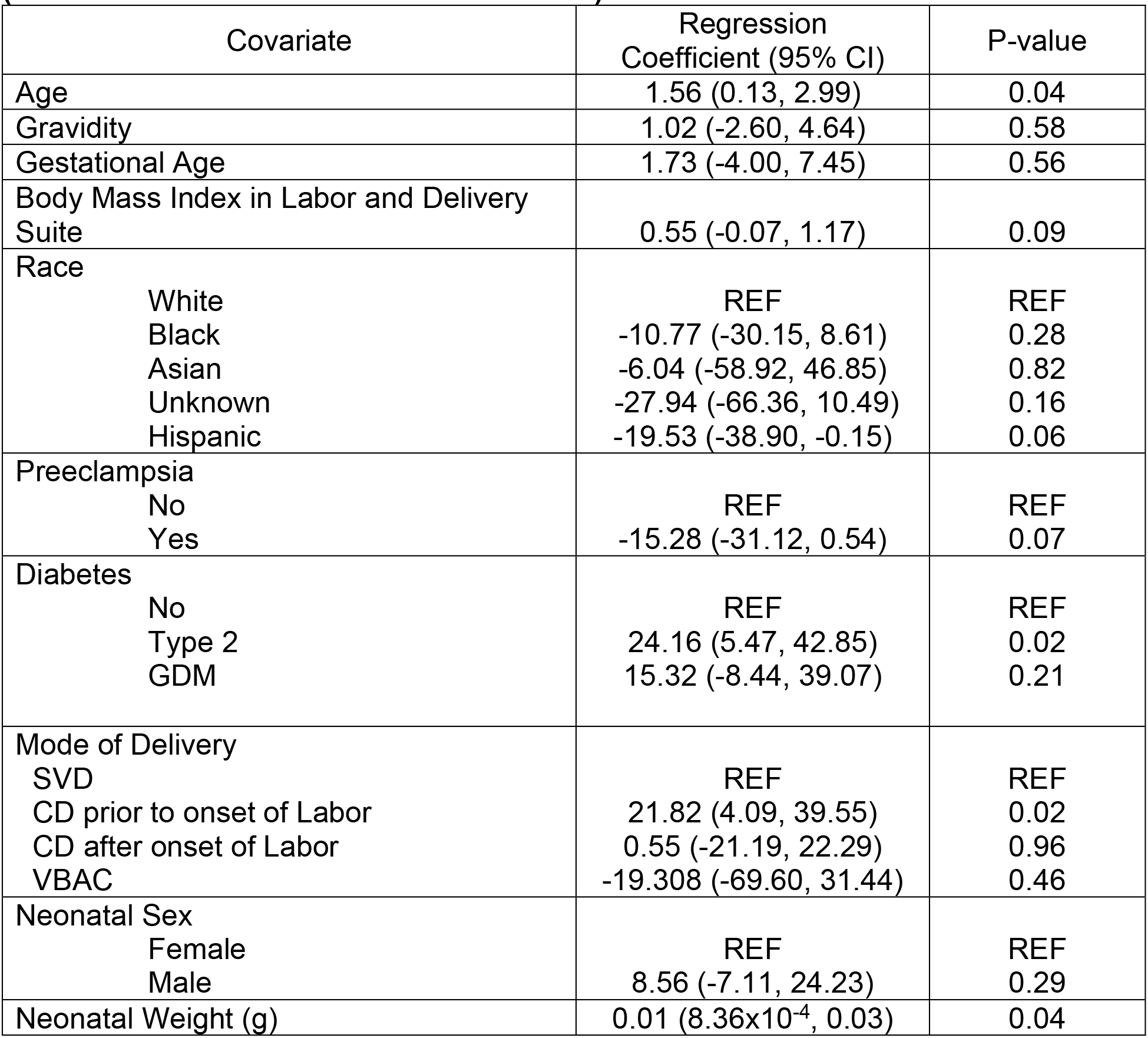
Regression coefficients from univariable analysis of OT-R protein band intensity (relative to internal reference individual T1)

V1aR protein levels were statistically significantly associated with mode of delivery, CD prior to onset of labor 20.36 (95% CI 10.58, 30.13) compared to SVD. See Table 9 for full results. No associations were found for race, neonatal sex, or gestational age between 35-42 weeks for any outcome.

**Table 9.**
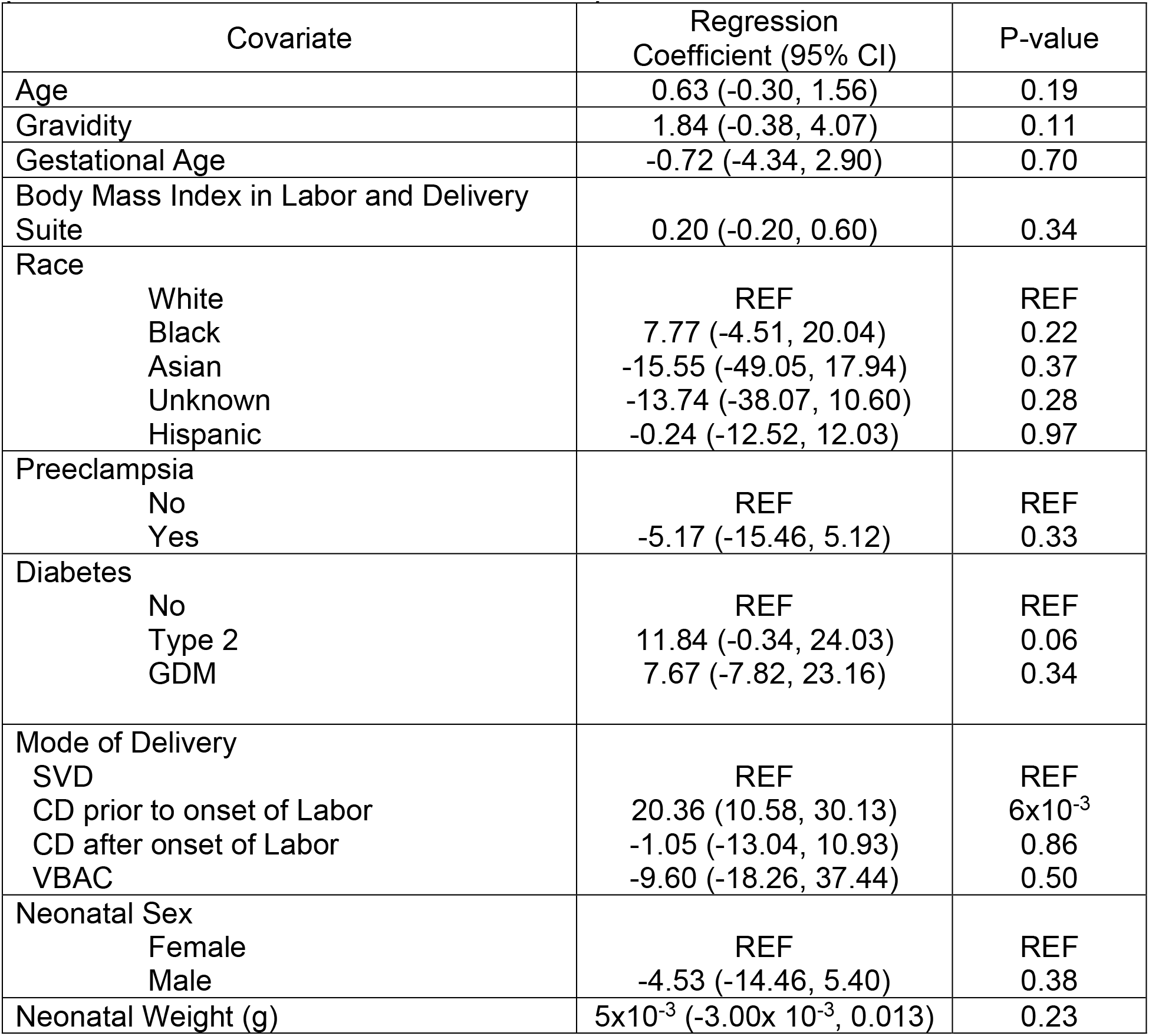
Regression coefficients from univariable analysis of V1aR protein band intensity (relative to internal reference individual T1)

### Multivariable analysis of protein levels

A multivariable model considered simultaneously all variables that were statistically significant in univariable analysis. MDMX protein levels showed a statistically significantly association with gravidity 7.19 (95% CI 1.24, 13.14), and preeclampsia -40.61 (95% CI -66.18, -15.04) as shown on Table 10. There were no significant associations for PICALM or OT-R as shown in Tables 11-12. The best fit model for V1aR included only an intercept (no covariate).

**Table 10.**
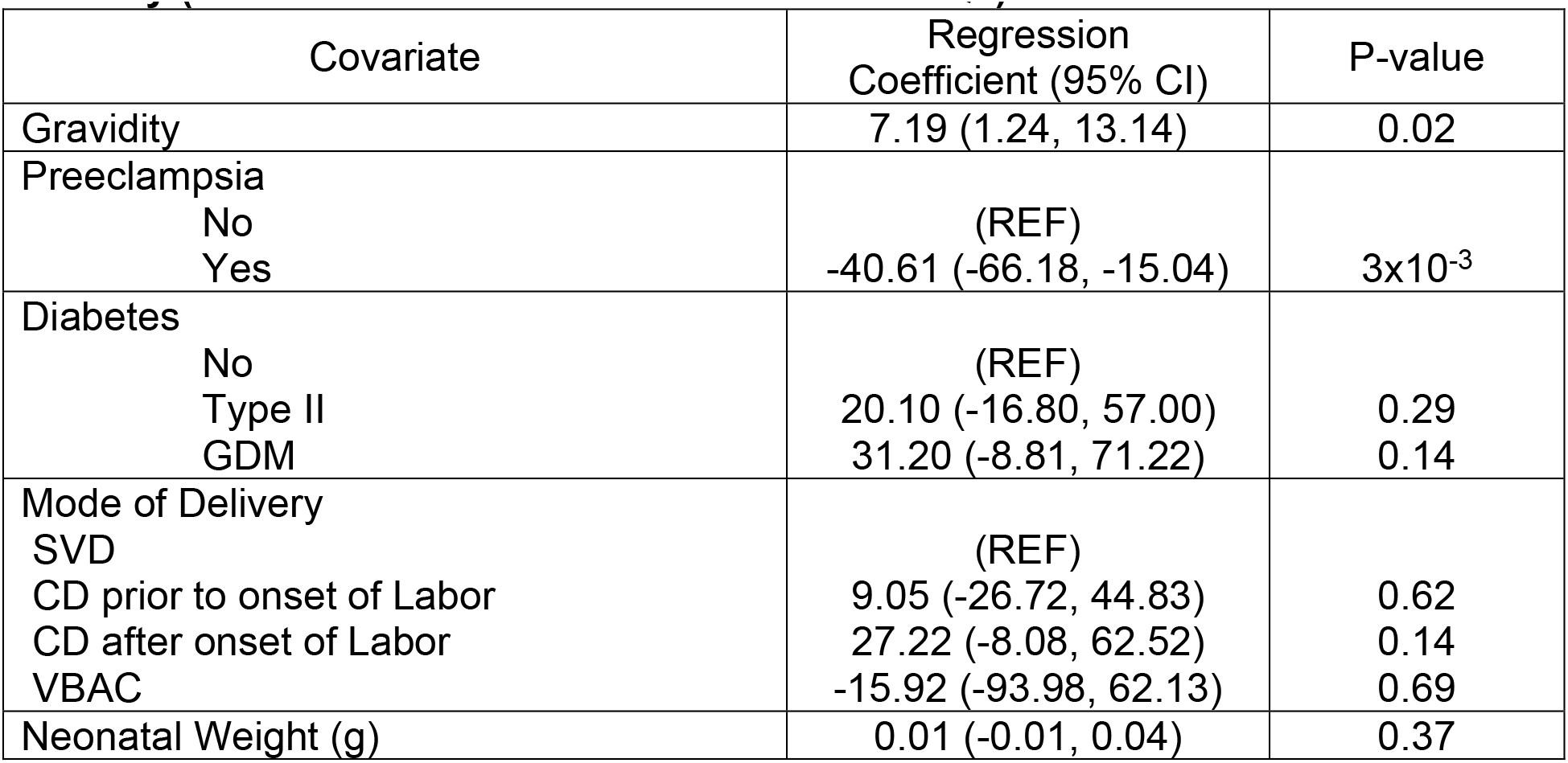
Regression coefficients from multivariable analysis of MDMX protein band intensity (relative to internal reference individual Q1)

| Covariate | Regression Coefficient (95% CI) | P-value |
| --- | --- | --- |
| Gravidity | 7.19 (1.24, 13.14) | 0.02 |
| Preeclampsia |  |  |
| No | (REF) |  |
| Yes | -40.61 (-66.18, -15.04) | 3x10<sup>-3</sup> |
| Diabetes |  |  |
| No | (REF) |  |
| Type II | 20.10 (-16.80, 57.00) | 0.29 |
| GDM | 31.20 (-8.81, 71.22) | 0.14 |
| Mode of Delivery |  |  |
| SVD | (REF) |  |
| CD prior to onset of Labor | 9.05 (-26.72, 44.83) | 0.62 |
| CD after onset of Labor | 27.22 (-8.08, 62.52) | 0.14 |
| VBAC | -15.92 (-93.98, 62.13) | 0.69 |
| Neonatal Weight (g) | 0.01 (-0.01, 0.04) | 0.37 |

**Table 11.**
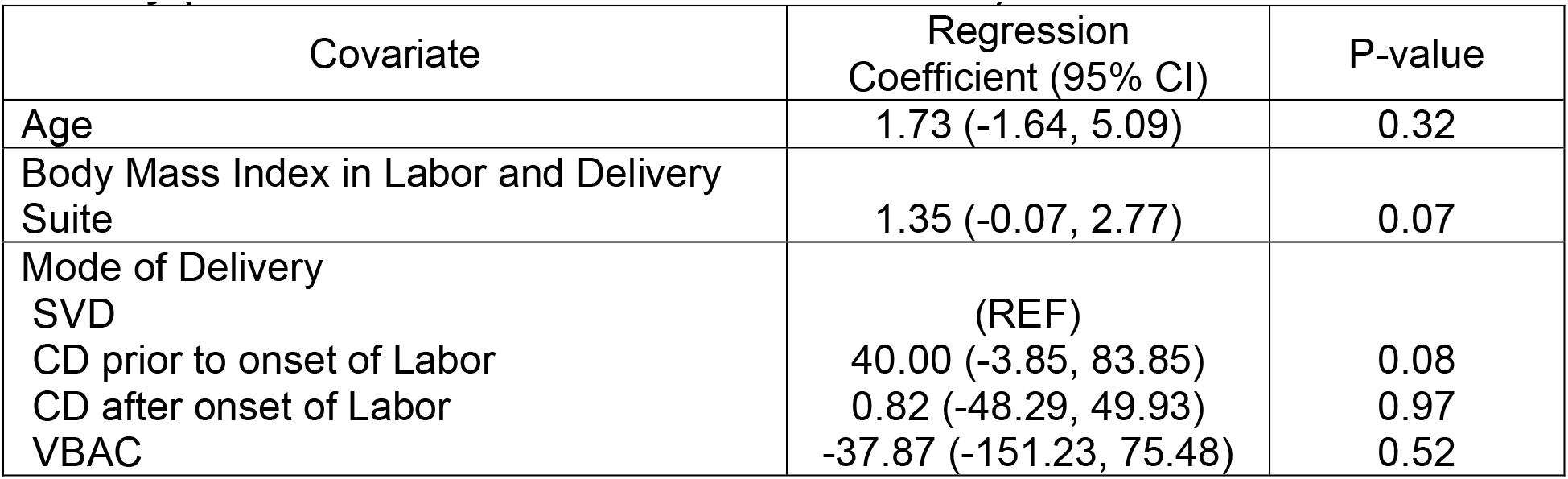
Regression coefficients from multivariable analysis of PICALM protein band intensity (relative to internal reference individual Q1)

| Covariate | Regression Coefficient (95% CI) | P-value |
| --- | --- | --- |
| Age | 1.73 (-1.64, 5.09) | 0.32 |
| Body Mass Index in Labor and Delivery Suite | 1.35 (-0.07, 2.77) | 0.07 |
| Mode of Delivery |  |  |
| SVD | (REF) |  |
| CD prior to onset of Labor | 40.00 (-3.85, 83.85) | 0.08 |
| CD after onset of Labor | 0.82 (-48.29, 49.93) | 0.97 |
| VBAC | -37.87 (-151.23, 75.48) | 0.52 |

**Table 12.** Regression coefficients from multivariable analysis of OT-R protein band intensity (relative to internal reference individual T1)

| Covariate | Regression Coefficient (95% CI) | P-value |
| --- | --- | --- |
| Age | 0.71 (-0.83, 2.35) | 0.39 |
| Diabetes |  |  |
| No | REF | REF |
| Type 2 | 14.0 (-11.14, 37.70) | 0.27 |
| GDM | 5.64 (-21.43, 32.00) | 0.68 |
| Mode of Delivery |  |  |
| SVD | REF | REF |
| CD prior to onset of Labor | 13.25 (-7.83, 34.33) | 0.23 |
| CD after onset of Labor | -1.04 (-23.92, 21.84) | 0.93 |
| VBAC | -0.17 (-68.44, 34.32) | 0.52 |
| Neonatal Weight (g) | $4.05 \times 10^{-4}$ (-0.02, 0.02) | 0.96 |

To account for the spread in the replicate measurements of MDMX, PICALM, OT- and V1aR (Table 5), we carried out the bootstrapping analysis shown in Tables 14-19. The statistical results were robust to bootstrapped resampling (n=1000) of data points included in the analyses.

**Table 13.** Regression coefficients from bootstrapped replicates of univariable analysis of MDMX protein band intensity (relative to internal reference individual Q1)

| Covariate | (95% quantiles of point estimate from bootstrapped replicates) | Percentage of replicates <0.05 (95% quantiles of P-value) |
| --- | --- | --- |
| Age | (0.43, 2.90) | 1.7% (0.05, 0.77) |
| Gravidity | (6.82, 12.15) | 95.5% ($8 \times 10^{-4}$, 0.06) |
| Gestational Age | (-6.88, 1.78) | 0% (0.25, 0.97) |
| Body Mass Index in Labor and Delivery Suite | (0.60, 1.50) | 12.6% (0.03, 0.34) |
| Race |  |  |
| White | (REF) |  |
| Black | (5.10, 39.4) | 0.9% (0.07, 0.80) |
| Asian | (-53.55, -6.43) | 0% (0.33, 0.91) |
| Unknown | (-30.86, 17.09) | 0% (0.46, 0.99) |
| Hispanic | (-19.69, 13.18) | 0% (0.32, 0.99) |
| Preeclampsia |  |  |
| No | (REF) |  |
| Yes | (-54.91, -28.15) | 91.2% ($9 \times 10^{-4}$, 0.08) |
| Diabetes |  |  |
| No | (REF) |  |
| Type II | (24.47, 60.41) | 60% ($2 \times 10^{-3}$, 0.23) |
| GDM | (24.83, 67.47) | 44.8% ($7 \times 10^{-3}$, 0.30) |
| Mode of Delivery |  |  |
| SVD | (REF) |  |
| CD prior to onset of Labor | (38.96, 67.36) | 98.6% ($5 \times 10^{-4}$, 0.04) |
| CD after onset of Labor | (17.16, 55.75) | 15.5% (0.18, 0.46) |
| VBAC | (-25.22, 55.19) | 0% (0.28, 0.99) |
| Neonate Sex |  |  |
| Female | (REF) |  |
| Male | (-10.62, 15.81) | 0% (0.32, 0.99) |
| Neonatal Weight (g) | (0.02, 0.05) | 85.8% ($3 \times 10^{-4}$, 0.14) |

**Table 14.** Regression coefficients from bootstrapped replicates of univariable analysis of PICALM protein band intensity (relative to internal reference individual Q1)

| Covariate | (95% quantiles of point estimate from bootstrapped replicates) | Percentage of replicates <0.05 (95% quantiles of P-value) |
| --- | --- | --- |
| Age | (2.19, 5.17) | 41% ($7.1 \times 10^{-3}$, 0.27) |
| Gravidity | (3.42, 11.80) | 22% (0.01, 0.46) |
| Gestational Age | (1.82, 11.72) | 0.1% (0.11, 0.82) |
| Body Mass Index in Labor and Delivery Suite | (1.41, 2.66) | 90% ($8 \times 10^{-4}$, 0.09) |
| Race |  |  |
| White | (REF) |  |
| Black | (-7.10, 40.07) | 0% (0.12, 0.97) |
| Asian | (-40.69, 111.00) | 0% (0.9, 0.99) |
| Unknown | (-96.07, -53.30) | 2% (0.05, 0.28) |
| Hispanic | (-64.85, -29.39) | 40% (0.01, 0.24) |
| Preeclampsia |  |  |
| No | (REF) |  |
| Yes | (-52.30, -18.88) | 20.3% (0.01, 0.37) |
| Diabetes |  |  |
| No | (REF) |  |
| Type II | (2.82, 45.52) | 0.6% (0.08, 0.91) |
| GDM | (23.35, 56.54) | 0.6% (0.08, 0.51) |
| Mode of Delivery |  |  |
| SVD | (REF) |  |
| CD prior to onset of Labor | (41.39, 79.89) | 88.5% ($9 \times 10^{-4}$, 0.08) |
| CD after onset of Labor | (-17.08, 31.02) | 0% (0.26, 0.99) |
| VBAC | (-82.60, -27.18) | 0% (0.22, 0.67) |
| Neonate Sex |  |  |
| Female | (REF) |  |
| Male | (3.42, 35.19) | 0.3% (0.10, 0.84) |
| Neonatal Weight (g) | (0.01, 0.05) | 35.1% ($3 \times 10^{-3}$, 0.40) |

**Table 15.**
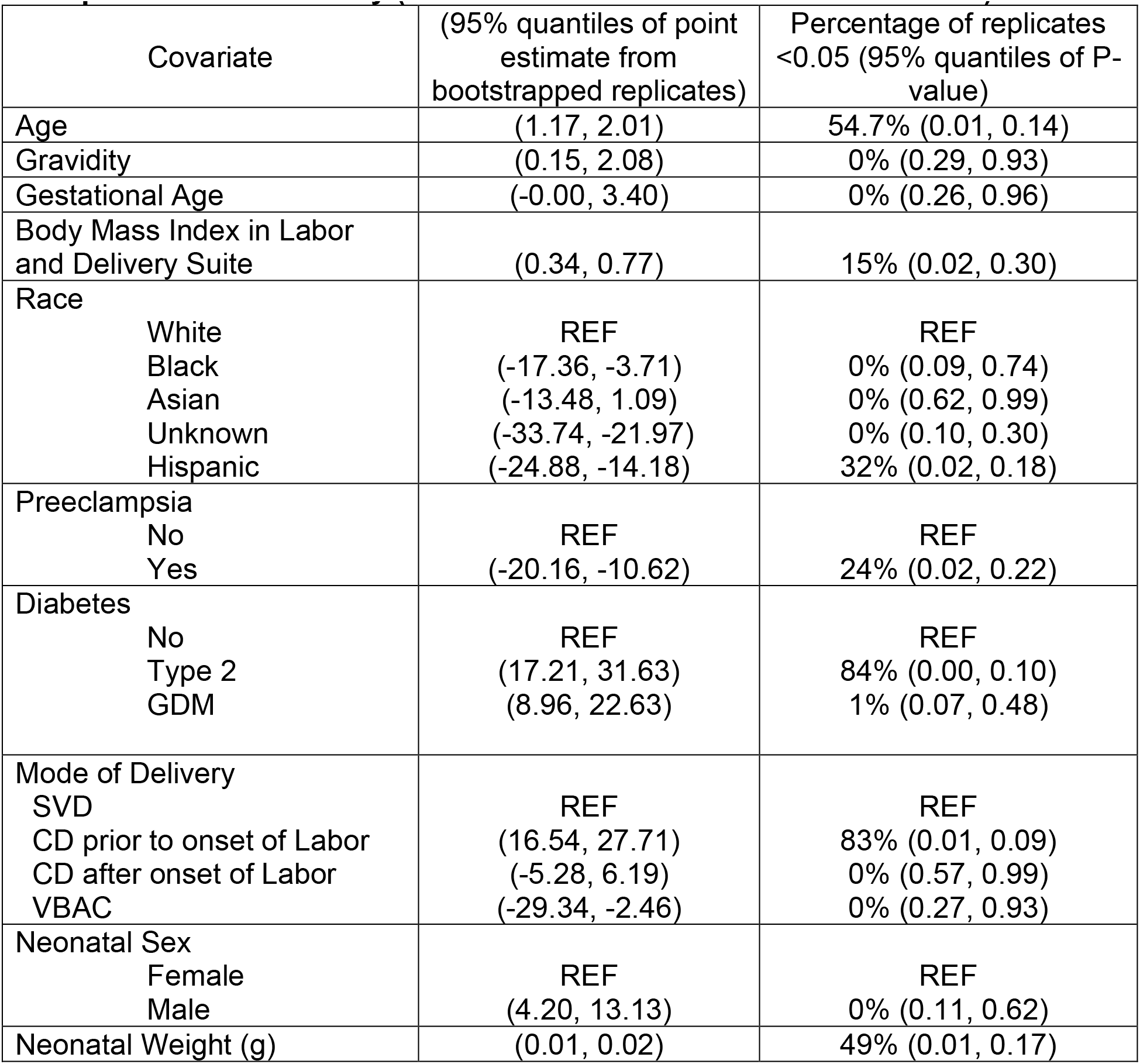
Regression coefficients from bootstrapped replicates of univariable analysis of OT-R protein band intensity (relative to internal reference individual T1)

| Covariate | (95% quantiles of point estimate from bootstrapped replicates) | Percentage of replicates <0.05 (95% quantiles of P-value) |
| --- | --- | --- |
| Age | (1.17, 2.01) | 54.7% (0.01, 0.14) |
| Gravidity | (0.15, 2.08) | 0% (0.29, 0.93) |
| Gestational Age | (-0.00, 3.40) | 0% (0.26, 0.96) |
| Body Mass Index in Labor and Delivery Suite | (0.34, 0.77) | 15% (0.02, 0.30) |
| Race |  |  |
| White | REF | REF |
| Black | (-17.36, -3.71) | 0% (0.09, 0.74) |
| Asian | (-13.48, 1.09) | 0% (0.62, 0.99) |
| Unknown | (-33.74, -21.97) | 0% (0.10, 0.30) |
| Hispanic | (-24.88, -14.18) | 32% (0.02, 0.18) |
| Preeclampsia |  |  |
| No | REF | REF |
| Yes | (-20.16, -10.62) | 24% (0.02, 0.22) |
| Diabetes |  |  |
| No | REF | REF |
| Type 2 | (17.21, 31.63) | 84% (0.00, 0.10) |
| GDM | (8.96, 22.63) | 1% (0.07, 0.48) |
| Mode of Delivery |  |  |
| SVD | REF | REF |
| CD prior to onset of Labor | (16.54, 27.71) | 83% (0.01, 0.09) |
| CD after onset of Labor | (-5.28, 6.19) | 0% (0.57, 0.99) |
| VBAC | (-29.34, -2.46) | 0% (0.27, 0.93) |
| Neonatal Sex |  |  |
| Female | REF | REF |
| Male | (4.20, 13.13) | 0% (0.11, 0.62) |
| Neonatal Weight (g) | (0.01, 0.02) | 49% (0.01, 0.17) |

**Table 16.** Regression coefficients from bootstrapped replicates of univariable analysis of V1aR protein band intensity (relative to internal reference individual T1)

| Covariate | (95% quantiles of point estimate from bootstrapped replicates) | Percentage of replicates <0.05 (95% quantiles of P-value) |
| --- | --- | --- |
| Age | (0.17, 1.12) | 6% (0.03, 0.74) |
| Gravidity | (0.48, 3.04) | 14% (0.02, 0.71) |
| Gestational Age | (-2.20, 0.98) | 0% (0.30, 0.98) |
| Body Mass Index in Labor and Delivery Suite | (-0.03, 0.43) | 2% (0.07, 0.93) |
| Race |  |  |
| White | REF | REF |
| Black | (1.07, 14.58) | 3% (0.04, 0.88) |
| Asian | (-32.74, -3.26) | 0% (0.09, 0.87) |
| Unknown | (-24.28, -1.85) | 0% (0.09, 0.89) |
| Hispanic | (-7.10, 7.06) | 0% (0.26, 0.99) |
| Preeclampsia |  |  |
| No | REF | REF |
| Yes | (-11.01, 0.40) | 2% (0.07, 0.94) |
| Diabetes |  |  |
| No | REF | REF |
| Type 2 | (5.78, 19.09) | 23% ($9 \times 10^{-3}$, 0.41) |
| GDM | (-4.01, 18.40) | 4% (0.04, 0.95) |
| Mode of Delivery |  |  |
| SVD | REF | REF |
| CD prior to onset of Labor | (14.50, 26.60) | 100% ($4 \times 10^{-5}$, $2 \times 10^{-2}$) |
| CD after onset of Labor | (-7.93, 5.70) | 0% (0.26, 0.99) |
| VBAC | (-7.09, 23.62) | 0% (0.16, 0.97) |
| Neonatal Sex |  |  |
| Female | REF | REF |
| Male | (-9.92, 0.86) | 0% (0.11, 0.93) |
| Neonatal Weight (g) | ($1.0 \times 10^{-3}$, $9.0 \times 10^{-3}$) | 2% (0.05, 0.82) |

**Table 17.**
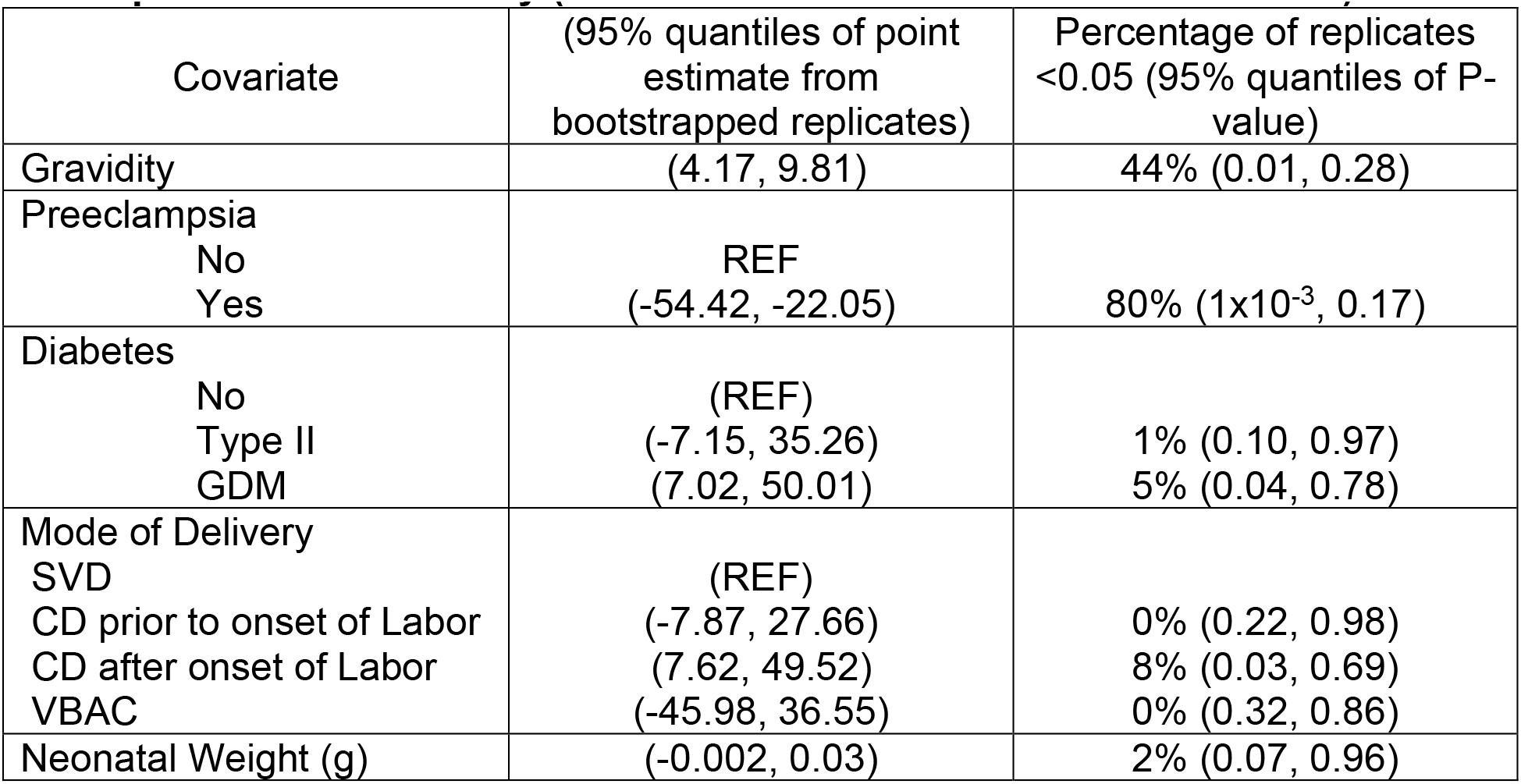
Regression coefficients from bootstrapped replicates of multivariable analysis of MDMX protein band intensity (relative to internal reference individual Q1)

**Table 18.** Regression coefficients from bootstrapped replicates of multivariable analysis of PICALM protein band intensity (relative to internal reference individual Q1)

| Covariate | (95% quantiles of point estimate from bootstrapped replicates) | Percentage of replicates <0.05 (95% quantiles of P-value) |
| --- | --- | --- |
| Age | (0.09, 3.35) | 1% (0.07, 0.93) |
| Body Mass Index in Labor and Delivery Suite | (0.55, 2.11) | 26% (0.008, 0.54) |
| Mode of Delivery |  |  |
| SVD | REF |  |
| CD prior to onset of Labor | (17.36, 62.80) | 20% (0.02, 0.48) |
| CD after onset of Labor | (-23.42, 26.23) | 0% (0.31, 0.99) |
| VBAC | (-70.17, -12.72) | 0% (0.29, 0.84) |

**Table 19.** Regression coefficients from bootstrapped replicates of multivariable analysis of OT-R protein band intensity (relative to internal reference individual T1)

| Covariate | (95% quantiles of point estimate from bootstrapped replicates) | Percentage of replicates <0.05 (95% quantiles of P-value) |
| --- | --- | --- |
| Age | (0.27, 1.17) | 0% (0.17, 0.76) |
| Diabetes |  |  |
| No | REF | REF |
| Type 2 | (6.09, 21.30) | 0% (0.10, 0.65) |
| GDM | (-1.66, 14.21) | 0% (0.32, 0.99) |
| Mode of Delivery |  |  |
| SVD | REF | REF |
| CD prior to onset of Labor | (7.33, 19.97) | 0% (0.09, 0.51) |
| CD after onset of Labor | (-6.97, 4.89) | 0% (0.55, 0.99) |
| VBAC | (-27.20, -0.44) | 0% (0.32, 0.98) |
| Neonatal Weight (g) | (-4x 10<sup>-3</sup>, 5x 10<sup>-3</sup>) | 0% (0.52, 0.99) |

We also looked at the correlation of MDMX, PICALM, OT-R and V1aR protein levels with each other and found significant correlations between PICALM and OT-R (0.72 95% CI 0.54, 0.84, p<2.7x10**^-8^**), PICALM and V1aR (0.41 95% CI 0.12, 0.63, p<0.006), and OT-R and V1aR (0.47 95% CI 0.20, 0.67, p<0.001). MDMX was not correlated with PICALM, OT-R or V1aR.

## Discussion

Mass spectrometry placed VEGFR2, MDMX and PICALM and their partners in the molecular landscape of chorionic villi of placentas at term. The co-immunoprecipitated proteins represent the most prevalent and stable complexes. IGEM provided a detailed map of VEGFR2, MDMX, PICALM, OT-R and V1aR in the villi, and hints about protein traffic, specifically, of placental exosomes transporting OT-R to the fetus. In future studies, IGEM can show if two proteins are within 10-20 Å of each other, with secondary antibodies labeled with gold particles of different size^42^, such as 6 nm and 12 nm, even though larger particles may occasionally eclipse the smaller ones.

A comprehensive statistical analysis of 44 placental extracts associated MDMX, PICALM, OT-R and V1aR protein levels with labor at term and a number of clinical parameters and gestational complications. Further studies may assign their binary protein interactions into networks of signaling partnerships among patient groups with normal gestation and gestational complications.^43^ The predominant expression of MDMX on fetal macrophages (HC)^37^ and the association of MDMX levels in the villi with most of the clinical characteristics we tested, suggest that MDMX has a central role in comorbidities, that originate from deficient trophoblast proliferation,^44^ and are linked to pre-existing conditions and environmental exposures.^45^

### MDMX immunoprecipitations

We were puzzled that the immunoprecipitated MDMX had a molecular mass of 50 kDa, instead of 75 kDa in the placental extracts (Fig. 1, B and E). A possible explanation was offered by the discovery^46^ that MDMX interacts with TRPM7, a bi-functional cation channel protein fused with a kinase domain. TRPM7 is a master regulator of the cellular balance of divalent cations, that mediates the uptake of Zn^2+^, Mg^2+^ and Ca^2+^,^47^ and senses oxidative stress to release Zn^2+^ from intracellular vesicles^48^. TRPM7 regulates MDMX levels by modulating Zn^2+^ concentration, and induces the formation of faster moving forms of MDMX on SDS-PAGE gels^46^. These forms depend on the channel function of TRPM7 and proteasomal degradation. We hypothesize that placental TRPM7, which is downregulated in preeclampsia,^49^ interacted with MDMX during the overnight incubation with the immunoprecipitation beads stripping MDMX of Zn^2+^. Proteasomal degradation was prevented by including MG-132 in our tissue-extraction and IP buffer. We missed detecting TRPM7 because it did not enter the top section of the 4- 12% Bis-Tris NuPAGE gels we used to fractionate the IP proteins with and submit the 25- 220 kDa strips for proteomic analysis (see Methods). TRPM7 has a theoretical mass of 213 kDa and pI 8.1, but on SDS-PAGE gels it is shown at 230 kDa and was reported at 245 kDa in BN electrophoresis.

### Autoantibody antigens and TRIM21

Although our patients did not show symptoms of autoimmune diseases, a few of the immunoprecipitated proteins listed on Table 20 were also among the autoantigens detected by Neiman *et al*. in healthy adults^50^, such as the extensively studied autoantigens TRIM21/Ro(SS-A) and PDC-E2, also an autoantigen in primary biliary cholangitis and other autoimmune diseases. PDC-E2 would be held on the beads as a partner of VEGFR2. TRIM21 is also extracted from intervillous maternal leucocytes based on their staining with TRIM21 (Fig. 2B). Maternal autoantigen-autoantibody complexes, with an affinity for proteins in the villous membrane fraction, would be trapped on the beads by Fc-receptor TRIM21 based on its high affinity for IgG^26^. Since TRIM21 has broad species specificity, it could bind to the sheep anti-rabbit Ab on the Dynabeads and the IP-bait, the anti-rabbit Ab the beads were charged with. If TRIM21 clusters on the beads remain catalytically active, they could, in principle, retain VEGFR2, MDMX and PICALM, as substrates of ubiquitin ligases, along with other proteins, many not *in vivo* TRIM21 substrates, detected by mass spectrometry. Consequently, TRIM21 can confound immunoprecipitations, performed according to our protocol, by retaining proteins unrelated to the bait-Ab. Based on the extensive TRIM21 literature, placental TRIM21 could participate in metabolic pathways as an E3 ubiquitin ligase,^51, 52^ and as Fc receptor defending against infections.

**Table 20.**
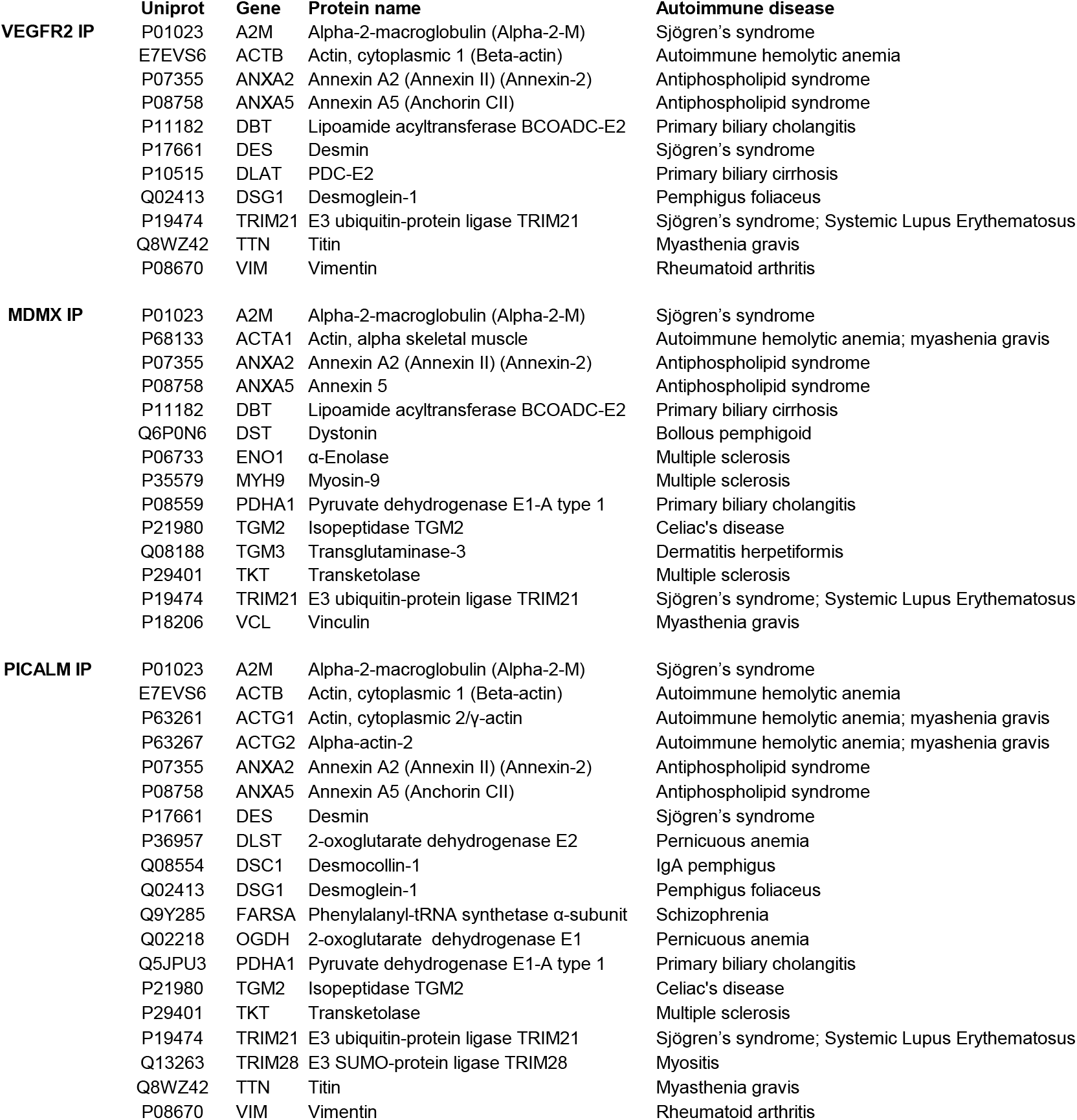
Autoantigens listed by Neiman et al. among the proteins in VEGFR2, MDMX, and PICALM immunoprecipitations (IP).

### Immunohistochemistry and immunogold electron microscopy

The distribution of VEGFR2, MDMX and PICALM in the villi was shown by IHC (Fig. 2) and by IGEM which also showed the distribution of OT-R and V1aR (Figures 3-7). VEGFR2 was detected in the nucleus, as previously reported^27, 53^, and in mitochondria, a novel observation to the best of our knowledge (Fig. 3, panels A, B and C). Translocation in the other direction, mitochondria to the nucleus, was shown for the entire pyruvate dehydrogenase complex to supply acetyl-CoA for histone acetylation^54^. VEGFR2 is distributed through relay networks in lipid rafts and endosomal trafficking^55^ and SUMOylation holds VEGFR2 at the traffic control Golgi apparatus^56^. Gradients in receptor concentration may exist along the placental vascular tree, as was shown for the purinergic P2Y2 receptor (pI=9.7)^57^. Distortion of receptor gradients in preeclampsia and diabetes could affect protein traffic and even expose proteins to degradation. Regarding the latter, it may be pertinent to consider the mechanism of action of estrogen receptor antagonists that decreased the estrogen receptor intra-nuclear mobility and subsequently induced its turnover^58^.

MDMX appeared in nuclear clusters (Fig. 4, A and B). PICALM was detected in nuclei (Fig. 5, B and C). The very basic OT-R and V1aR were also detected (Figures 6 and 7). Surprisingly, clusters of V1aR became clearly visible on a fetal RBC after increasing gamma (Fig. 7, C). Since vasopressin receptors participate in erythropoiesis^59^ their presence on RBC was not very surprising. We hypothesize that in addition to non-traditional functions, such as critical immune sensors,^60^ RBC transport V1aR to the endothelial cells of the villi^25^.

PICALM and OT-R were detected on endothelial cell projections into the fetal lumen (Figures 5A, 6A) where OT-R clusters were also observed (Fig. 6, A and B) matching the dimensions of microvesicles-exosomes^10, 11^. Although we cannot prove that OT-R clusters are inside exosomes with the characteristic bi-lipid membranes, as seen in standard EM cellular morphology, we propose that fetal exosomes carry to the fetus the OT-R and other proteins produced in the placenta. A limitation of IGEM, from the exosome perspective, is that bi-lipid layers are greatly impaired during the processing of tissues because OsO4 is omitted in a post-fixation step used to increase the visualization of such membranes. OsO4 reacts strongly with lipid complexes, and while it enhances contrast for standard EM cell morphological imaging, it can oxidize many antigen epitopes in IGEM. Fig. 8 shows representative intraluminal vesicles^9^ seen in the fetal capillary of the placenta sample that was processed with the OsO4 post-fixational step included. Membranes are more clearly defined here, compared to the IGEM sample, representing exosomes or microvesicles^9^ in the fetal lumen (Fig. 8, A and B). We noticed similarities between our IGEM Figures 5 and 6 and transmission electron microscopy images in a study at two polluted cities in Mexico showing that environmental nanoparticles of Fe, Ti, Cu, Hg and Sn accumulated in HC and endothelial cells of chorionic villi sampled at term^61^.

### Statistical analysis of protein levels and clinical presentations

Our search for the function of the newly detected placental membrane proteins MDMX, PICALM, and of OT- R and V1aR, was accelerated by a statistical analysis of their protein levels, which revealed numerous associations with clinical characteristics of the 44 patients. MDMX, located predominantly in HC (Fig. 2C), took center stage based on its association with most of the clinical characteristics we tested. Although many of our statistical associations from 44 placentas are highly unlikely to be observed by chance, our conclusions could change by testing a larger number of placenta samples and integrating, after further research, protein isoforms and post-translational modifications of the newly detected proteins. But to what extent did proteins from intervillous blood, reflecting also the maternal immune system,^45^ contribute to the observed clinical associations? An initial answer was provided for MDMX by the statistical analyses showing that tissue-resident HC contributed most extensively since they predominantly express MDMX (Fig. 2).

### Diabetes

In diabetic patients, higher MDMX levels (Table 6) seem to correspond to the higher number of cells stained with CD163^62^, a marker of HC^39^. It is not known if MDMX upregulation and higher HC numbers mark a metabolic shift towards diabetes. A diabetic atherosclerosis rat model showed upregulation of MDMX mRNA and protein levels^63^. MDMX was shown in cell cultures to serve as a nutrient sensor by inhibiting mTORC1^64^. HC could facilitate nutrient transportation in the villous stroma^34^ linking placental MDMX to nutrient levels and metabolic networks. OT-R levels are higher in Type 2 diabetes (Table 8), and there is an association of *OXTR* variants with insulin sensitivity and Type 2 diabetes^65^.

### Preeclampsia

Villous trophoblasts in pregnancies complicated by preeclampsia had higher p53 and lower MDM2 levels ^18^. Although MDMX was not measured, we believe it would have been decreased also. Lower MDMX levels (Tables 6, 10) could reflect the decreased numbers of HC in preeclampsia^40, 41^. What can cause the decrease of HC? In cancer, abnormal vascularity disrupts the penetration of immune cells,^66^ and their function is hindered by the tumor microenvironment^67, 68^. Here, defective placental vascularity and increased placental stiffness^69^, both changing the architecture of the villi, could have an adverse effect on HC levels. In preeclampsia, placental stiffness could be increased by higher levels of tissue transglutaminase/*TGM2* (Tables 3, 4) which catalyzes protein crosslinking^32^. MDMX could be involved in the reprogramming of fetal macrophages (HC) during preeclampsia and other gestational complications and labor. Drawing from the breast cancer literature^70^, downregulation of placental MDMX in preeclampsia could be linked to estrogen receptor-α^71^ and lower estrogen levels^72, 73^.

In preeclampsia large quantities of magnesium sulfate are prescribed to prevent seizures. A potentially relevant study of a blood-brain barrier (BBB) model of primary endothelial cells from human brain^74^, showed that TRPM7 mediated the entry of extracellular Mg^2+^ into cells and that high Mg^2+^ levels speeded up the clearance of Aβ to the blood side via BBB transcytosis and upregulation of PICALM and LRP-1 that is also expressed in HC^75^. It was shown recently that TRPM7 kinase activity induces amyloid-β degradation and clearance^76^. Among other mechanisms, the accumulation of placental Aβ^21^ could be related to TRPM7 and limiting PICALM-dependent transcytosis.

### Gravidity

Univariable and multivariable regression analysis found a significant association of gravidity with MDMX levels (Tables 6, 10). Gravidity was recently associated with HC levels determined by CD163 immunostaining^41^. While multigravida women have shorter telomeres and increased DNA methylation age,^77^ a role of p53 and MDMX on telomere length is not established in normal human tissues, to the best of our knowledge. DNA methylation age is associated with MDMX and prenatal smoke exposure^78^. The gravidity-MDMX association, which registers in the villi, could reflect the cost of multiple pregnancies to the mother^79^.

### PICALM, OT-R and V1aR

The association of placental PICALM levels with BMI (Table 7) is intriguing because the placenta functions in the absence of adipocytes. A potential precedent for such association was provided by a study on gastric bypass surgery where PICALM mRNA levels in the blood were decreased after a significant drop of BMI^80^. Furthermore, among the Alzheimer’s risk nucleotide polymorphisms (SNP), a PICALM SNP is associated with obesity^81^.

The strong correlation of protein levels of PICALM with OT-R (p<2.7x10**^-8^**), PICALM with V1aR (p<0.006), and OT-R with V1aR (p<0.001) point to fundamental interactions by yet unknown mechanisms. MDMX was not correlated with PICALM, OT-R or V1aR.

Downregulation of OT-R in spontaneous vaginal deliveries (SVD) relative to CD prior to the onset of labor (Table 8), would be consistent with the higher oxytocin levels in SVD than CD. It is not known to what extend extravillous blood cells and exosomes may have supplied OT-R, and V1aR, detected in placental membrane extracts. Nevertheless, IGEM images of the chorionic villi support the hypothesis that the lipid-covered exosomes carry a cargo of the very basic, magnesium-dependent OT-R across the blood-brain barrier to interact also with fetal microglia^82–84^, the resident immune cells of the brain^85^. OT- R protein levels in venous and arterial blood from the umbilical cord may reveal differences in bi-directional OT-R traffic in normal, preterm and postterm deliveries. In that case, OT- R protein levels and epigenetic modifications^86^ may lead to therapeutic interventions with exosomes^87^ prepared, ideally, with the transmembrane orientation^12^ of OT-R in villous exosomes.

### Labor

The decrease of MDMX in the membrane fraction of chorionic villi in SVD vs. CD prior to the onset of labor (Table 6), could be related to the decrease of immune cells in peripheral maternal blood with approaching labor^88^, and the molecular signature of the “immune clock”.^45, 89^ Lower MDMX levels could also indicate a molecular switch in late gestation, locally, to energy-preserving measures or other mechanisms involving uterine macrophages^90^. MDMX, PICALM, OT-R and V1aR could be components of the fetal and maternal immune system during pregnancy and as the fetus develops immunity in preparation for birth.^91, 92^ Concomitant downregulation of MDMX, PICALM, OT-R and V1aR occurred only in SVD compared to CD prior to the onset of labor (Tables 6-9) signifying a potential convergence of metabolic networks to prepare the uterus for the final act in pregnancy, delivery of the fetus.

## Materials and methods

### Human Subjects

All patients signed consent forms prior to their inclusion in the study according to the protocol approved by the Institutional Review Board of the University of South Florida and Tampa General Hospital. Placental samples were obtained from 44 patients with singleton pregnancies, aged 19-40 and gestational ages ranging from 35-42 weeks based on the dating criteria from the American College of Obstetricians and Gynecologists (ACOG) (https://pubmed.ncbi.nlm.nih.gov/28426621/) for women who delivered vaginally or by cesarean delivery (CD), between 7 AM to 4 PM, based on obstetric indications (Table 1). Patients were excluded if they had active viral infections or fetal growth restriction, defined as estimated fetal weight of <10^th^% per the Hadlock growth curve. Preeclampsia was diagnosed according to the criteria established in 2013 by ACOG (DOI: <u>10.1097/01.AOG.0000437382.03963.88)</u>. Low risk pregnancy was defined by the absence of maternal co-morbidities including chronic hypertension defined as hypertension prior to pregnancy or systolic blood pressure ≥140 mm Hg or diastolic blood pressure ≥90 mm Hg prior to 20 weeks’ gestation, pregestational or gestational diabetes, smoking, renal disease, and autoimmune disease. Patients with preeclampsia with severe features defined by the 2013 ACOG Guidelines received magnesium sulfate for seizure prophylaxis during induction of labor or CD through 24*h* postpartum. The demographic Table 21 shows that 91% of study participants were 18-34 years old and 75% were multiparous. Most participants (91%) were ≥37 weeks’ gestation. There was no risk of bias in the selection of placentas which was at random. Sample/patient IDs (e.g., H-1, J-1, etc.) are not known to anyone outside the research group, and are assigned identifiers not using any personally identifiable information.

**Table 21.** Patient demographics.

Patient demographics
| Clinical characteristics | Study subjects (n=44) | % of n |
| --- | --- | --- |
| Maternal Age |  |  |
| 18-34 | 40 | 91% |
| ≥35 | 4 | 9% |
| BMI |  |  |
| Underweight (<18.5) | 0 | 0% |
| Normal (18.5-24.9) | 3 | 7% |
| Overweight (25-29.9) | 9 | 20% |
| Obesity I (30-34.9) | 12 | 27% |
| Obesity II (35-35.9) | 5 | 11% |
| Extreme Obesity (≥40) | 15 | 34% |
| Parity |  |  |
| Nulliparous | 11 | 25% |
| Multiparous | 33 | 75% |
| Gestational Age at Delivery |  |  |
| < 37 weeks | 4 | 9% |
| ≥ 37 weeks | 40 | 91% |
| Race |  |  |
| White | 17 | 39% |
| African American | 12 | 27% |
| Hispanic | 12 | 27% |
| Other | 3 | 7% |
| Diabetes | 14 | 32% |
| Type 1 | 0 | 0% |
| Type 2 | 9 | 20% |
| Gestational | 5 | 11% |
| Chronic Hypertension | 6 | 14% |
| Normotensive | 28 | 64% |
| Preeclampsia | 16 | 36% |
| Preeclampsia and Diabetes | 4 | 9% |

### Reagents

Most of the reagents were purchased from Thermo Fisher Scientific (Waltham, MA, USA), MMP-200 metalloprotease inhibitor III, proteasome inhibitor MG-132, protease inhibitor cocktail III, marimastat, Hammarsten-grade casein, detergent ASB-14, Novagen S-protein HRP conjugate 69047-3, Novex 4-12% BT SDS-PAGE gels, CL-XPosure film, magnetic Dynabeads M-280 charged with sheep anti-rabbit antibodies, Maxisorp Nunc C-bottom 8-well strips, protein assay Pierce BCA 562 nm kit 23225 (Pierce, Rockford, IL), and Protein Perfect HRP MW markers 69079-3/ Millipore-Calbiochem (Billerica, MA). Additional reagents are provided in the sections below.

### Antibodies (Protein target/vendor)

VEGFR2 (55B11) no. 2479/Cell Signaling (Danvers, MA), MDMX A300-287A/Bethyl Laboratories (Montgomery, TX), PDC-E2 SC-365276/Santa Cruz Biotechnology (Santa Cruz, CA), PICALM Sigma Prestige HPA019053, OT-R ABN1735/Millipore (Temecula, CA), V1aR MBS176788/MyBiosource (San Diego, CA), CD163 MA5-33091/InVitrogen-Thermo Fisher Scientific, TRIM21 Novus NBP1-33548 (Centennial, CO), Alexa-488 (green) and Alexa-64 (red) goat anti-rabbit/mouse conjugated secondary antibodies/Life Technologies (Eugene, OR), immunogold donkey-anti-rabbit secondary antibodies 25702 (6 nm) and 25705 (10 nm) (Electron Microscopy Sciences (Hatfield, PA).

### Placenta tissue collection, homogenization, and protein extraction with ASB-14

Within 15 min of the delivery of the placenta^3^,^0^samples were obtained from the fetal side of the placental bed at 4-5 cm from the umbilical cord and were placed on dry ice and then stored at -80°C. Tissues were also fixed in formalin for IHC or in paraformaldehyde-tannic acid for IGEM. Frozen tissue samples were finely cut and placed, at 150 mg per mL, in homogenization buffer containing 50 mM sodium phosphate pH 7.6, 50 mM NaCl, 50 μM sodium o-vanadate, 10 mM NaF, 20 μM proteasome inhibitor MG-132, 10 μM marimastat and 6 μL protease inhibitor cocktail-III per mL. After homogenization at 4^0^C by three 15s bursts of a homogenizer with one-minute cooling in between, the homogenates were centrifuged for 1h at 100,000*g*. On top of the pellets was a thin pink fluffy layer, presumably of RBC membranes, which was included when the pellets were suspended in homogenization buffer containing 1% ASB-14 and then extracted overnight, due to logistical reasons, at 6-8^0^C by end-over-end rotation. The zwitterionic detergent ASB-14 was superior in solubilizing membrane proteins to CHAPSO or n-octyl β-D-maltoside. After centrifugation at 100,000*g* for 1h, the supernatant solution was used in immunoprecipitations and western blots. We refrained from washing the membrane pellets in order to capture weak protein complexes. Protein levels of the membrane extracts were determined in quadruplicate at two dilutions with the Pierce BCA 562 nm kit. Final yield: 10-15 mg of detergent-extracted proteins per g wet tissue. To determine the efficiency of protein extraction by ASB-14, the post-ASB-14 100,000*g* pellets were solubilized in SDS-DTT at 95^0^C and then probed in western blots. Extraction yield was 100% for VEGFR2, MDMX and PICALM, 86% for PDC-E2 and 64% for TOMM20, a marker for inner mitochondrial membranes.

### Peptide synthesis to validate the OT-R and V1aR antibodies

Fig. 10 shows OT-R peptides EGNRTAGPPRRNEA (within the OT-R immunogen range given for this antibody), AEAPEGAAAGDGGRVA (outside the OT-R immunogen range) and V1aR immunogen peptide HPLKTLQQPARRSRLMIAA to validate the OT-R Millipore ABN1735 (0.5 μg/μL) and V1aR MyBioSourse MBS176788 (0.5 μg/μL) antibodies. The peptides were synthesized on 100 mg Rink Amide-MBHA resin (0.65 mmol/g) at room temperature under air using standard solid phase peptide synthesis protocol.

### Validation method for OT-R and V1aR antibodies

Maxisorp Nunc C-bottom strips were coated overnight at 4^0^C with the indicated amounts of synthetic peptides (Fig. 10) dissolved in sodium carbonate buffer pH 9.6^93^. After blocking, as in western blot experiments, the OT-R and V1aR antibodies were added for 2h at room temperature at the indicated dilution followed by the secondary anti-rabbit antibody conjugated with horseradish peroxidase. Detection of bound OT-R or V1aR antibodies was conducted by ECL on films exposed for the indicated times. The OT-R and V1aR antibodies bound only to peptide sequences used to raise them.

### Immunoprecipitations

Each immunoprecipitation of the 4-6 placental extracts, shown in Table 3-5, was carried out in one batch. Half mL of pellicular support M-280 Dynabeads was washed in homogenization buffer containing 1% ASB-14 and charged separately at room temperature for 2h with 0.9 μg VEGFR2, 15 μg MDMX or 7.5 μg PICALM antibody. Three mg of ASB14-extracted placental membranes from each placenta were mixed separately with the beads, overnight for logistical reasons, at 6-8^0^C. Most proteins adhering to the beads were removed by washing four times with homogenization buffer containing 1% ASB-14 and once with detergent-free buffer. Bound proteins were eluted at 95^0^C with 100 mM Tris-HCl pH 7.6, 4% SDS (w/v) and 100 mM DTT. Eluted proteins were fractionated on 4-12% Bis-Tris NuPAGE gels. After Coomassie blue staining, gel strips from 25-220 kDa were submitted for mass spectrometric analysis.

### Mass spectrometry

Samples were submitted and analyzed in a double-blinded fashion. Immunoprecipitated proteins were digested with Trypsin/Lys-C overnight at 37^0^ C as described^94^. Each group of immunoprecipitated samples was analyzed with a new trap and HPLC column. Peptides were dried in a vacuum concentrator and resuspended in 0.1% formic acid for LC-MS/MS analysis. Peptides were separated with a C18 reversed-phase-HPLC column on an Ultimate3000 UHPLC with a 60-min gradient and analyzed on a Q-Exactive Plus using data-dependent acquisition. Raw data files were processed in MaxQuant (www.maxquant.org) and searched against the UniprotKB human protein sequence database. Search parameters included constant modification of cysteine by carbamidomethylation and the variable modification, methionine oxidation. Proteins were identified using the filtering criteria of 1% protein and peptide false discovery rate.

### Western blot analysis

25 μg of protein from placental extracts were resolved on 4-12% Bis-Tris NuPAGE gels, transferred to PVDF membranes and blocked overnight at room temperature in 1% Hammarsten-grade casein or 5% Difco skimmed milk powder. MDMX, PICALM, OT-R or V1aR primary antibody dilutions ranged from 1:800 to 1:10000. PVDF membranes incubated with ECL reagents were exposed to film for 1-20s. Films were scanned in the transmittance mode (Epson Perfection 3200 Photo, Epson America, Los Alamitos, CA, USA). The intensity of the top band (Fig. 9) was measured with Li-Cor Image Studio Lite software (Lincoln, Nebraska, USA) and chosen from a film exposed at the linear portion of the correlation between band intensity and film exposure time before films were overexposed. In the statistical analyses we used the mean intensity from 3.6 blots, on the average, and relative to an internal control sample Q1 or V1 (Table 5).

### Immunohistochemistry

4 µm formalin-fixed paraffin embedded tissue sections were processed for IHC and stained with VEGFR2 (1:50), MDMX (1:500) or PICALM (1:1000) antibodies. Slides were exposed with diaminobenzidine tetrahydrochloride dehydrate as a chromogen and counterstained with hematoxylin before permanent mounting.

### Immunogold electron microscopy

Tissue samples were promptly fixed in buffered 4% paraformaldehyde and kept overnight at 4^0^C and then immersed in 0.75% buffered solution of tannic acid for 2h at room temperature. Samples were rinsed in filtered glass-distilled water, dehydrated through a series of increasing ethanol concentration, infiltrated, and embedded in LRW acrylic resin. Selected blocks were sectioned at 90-100 nm and mounted on formvar coated copper grids. The immunogold reaction was performed by incubating grids in standard blocking solution for 30 min at room temperature and transferring to either 1:100 MDMX rabbit Ab, 1:400 PICALM rabbit Ab, 1:100 VEGFR2 rabbit Ab or 1:600 OT-R or V1aR rabbit Ab for an overnight incubation at 4^0^C. After washing in PBS, gold-conjugated donkey anti-rabbit Ab 25703 (Electron Microscopy Sciences, Hatfield, PA), was applied at 1:50 dilution for 90 min at room temperature. The diameter of the gold particles (Electron Microscopy Sciences, Hatfield, PA) was 6 nm in VEGFR2 and MDMX and 10 nm in PICALM and OT-R experiments. Excess secondary antibody was removed by washing with glass-distilled water and the grids were further treated with 2% aqueous uranyl acetate added for contrast. In control experiments excluding the primary antibody, 1-2 gold particles were detected throughout the grids. In morphological analysis by transmission electron microscopy, tissues were fixed in 2% OsO4, after 2.5% glutaraldehyde fixation and prior to the dehydration and resin infiltration steps. Images were obtained with a JEM-1400 transmission electron microscope (JEOL, Peabody, MA) and Orius-832 camera (Gatan Inc., Pleasanton, CA).

### Whole mount immunofluorescence (WMIF)

Small samples (2x2x2 mm) of placental tissues ,were fixed in 1 mL 90% methanol for 2*h* at 4°C according to Bushway et al.^95^ with modifications. After washing with PBS, the samples were blocked for 2h at 4^0^C with 1% Hammarsten-grade casein in PBS containing 0.02% Thimerosal. After washing with 0.5% Casein in PBS, antibodies to VEGFR2 and PDC-E2 were added at 1:400 dilution and incubated overnight at 4^0^C, followed by washing with 0.5% Casein-PBS containing 0.3% Triton-X and incubation with the secondary antibodies at 1:100 dilutions. WMIF slides were observed with a Leica TCS SP5 AOBS laser scanning confocal microscope through a 40X/1.3NA Plan Apochromat oil immersion objective lens (Leica Microsystems CMS GmbH, Germany). 405 Diode, Argon 488 and He-Ne 647 laser lines were applied to excite the samples and tunable emissions were used to minimize crosstalk between fluorochromes. Images were captured with photomultiplier detectors and prepared with the LAS AF software version 2.7 (Leica Microsystems CMS GmbH, Germany). A 3D projected image was created by Z-stack of images taken at 1.0-0.5-micron intervals to create a movie by Imaris version 7.6 (Bitplane AG, Switzerland) ( video).

### Statistical analysis

The protein levels of MDMX, PICALM, OT-R and V1aR were analyzed for univariable and multivariable associations by linear regression using the R statistical program (https://www.r-project.org, 4.1.3). Parsimonious multivariable models were selected using Akaike’s Information Criteria with a stepwise selection procedure (StepAIC in the MASS package of R) and were run with covariates that were identified as statistically significant in univariable analysis. A p-value of less than 0.05 was considered statistically significant. Violin plots were made by R statistical program, and data Figures and graphs were assembled using Fiji ImageJ, GraphPad Prism 9.4.0, Adobe Illustrator and Windows Office365 software. To test the robustness of results to the specific datapoints included in the analysis, bootstrapped resampling (with replacement) was used to generate 1000 replicate datasets, upon which regression analyses were redone. Results of bootstrapped resampled analyses were summarized and compared to the full dataset to characterize consistency of point estimates and patterns of statistical significance. Pearson correlation coefficients were used to characterize correlation of protein measurements with each other.

## Acknowledgements

This study was supported by the Department of Obstetrics and Gynecology at the University of South Florida and research funds from Tampa General Hospital (AEA) and the Teasley Foundation (TJR). The WMIF images were obtained by the Analytic Microscopy Core Facility at the H. Lee Moffitt Cancer Center & Research Institute, an NCI designated Comprehensive Cancer Center (P30-CA076292). We thank Umit A. Kayisli for advice on IHC, and Santo V. Nicosia and Greg Arsenis for reading the manuscript and comments.

## Author contributions

JCMT conceived the study, performed most wet chemistry experiments, and wrote the first draft of the manuscript in consultation with VNU and RRM. SJH, RGS, and EPN recruited patients, collected, and interpreted clinical data in consultation with TSS, TJR, AEA, and MLA. DC performed proteomic analyses. MO assisted in the analysis of western blot data and confocal and WMIF experiments. SA and NS performed IHC experiments. AG performed and interpreted IGEM. PS and JC synthesized OT-R and V1aR peptides. DATC performed analyses and statistical data presentation. All authors critically read and approved the manuscript.

## Data availability

The video of the chorionic villi is deposited at https://nam04.safelinks.protection.outlook.com/?url=https%3A%2F%2Fzenodo.org%2Frecord%2F8160169%3Ftoken%3DeyJhbGciOiJIUzUxMiIsImV4cCI6MTY5MjMwOTU5OSwiaWF0IjoxNjg5Njk3NDQzfQ.eyJkYXRhIjp7InJlY2lkIjo4MTYwMTY5fSwiaWQiOjM1ODEzLCJybmQiOiJjMTlkOTM2NCJ9.2OSYKMRxBpqO7afBKem_S8HdQJgmbtnahLFTgJ_5hoiK3wxcbB2fvuyqEPJ0PeTtF_r9f9Zuocr6LcfXPs2-gw&data=05%7C01%7Ctsibris%40usf.edu%7C02564875fb61480dab4f08db87ae6fdb%7C741bf7dee2e546df8d6782607df9deaa%7C0%7C0%7C638252955485880125%7CUnknown%7CTWFpbGZsb3d8eyJWIjoiMC4wLjAwMDAiLCJQIjoiV2luMzIiLCJBTiI6Ik1haWwiLCJXVCI6Mn0%3D%7C3000%7C%7C%7C&sdata=iukZjlFOvoIiQAdfd566alnMH33jLMz3TV%2BDqPv5Trk%3D&reserved=0.

All MS raw files and corresponding results files will be deposited to the ProteomeXchange Consortium via the PRIDE partner repository.

